# Investigating the association between fasting insulin, erythrocytosis and HbA1c through Mendelian randomization and observational analyses

**DOI:** 10.1101/2022.12.21.22283813

**Authors:** Anthony Nguyen, Rana Khafagy, Habiba Hashemy, Kevin H.M. Kuo, Delnaz Roshandel, Andrew D. Paterson, Satya Dash

## Abstract

**Background:** Insulin resistance (IR)/hyperinsulinemia (HI), are early abnormalities in the etiology of prediabetes (preT2D) and type 2 diabetes (T2D). IR/HI also associate with increased erythrocytosis. Hemoglobin A1c (HbA1c) is commonly used to diagnose and monitor preT2D/T2D, but can be influenced by erythrocytosis independent of glycemi.

**Methods:** We undertook bidirectional Mendelian randomization (MR), in individuals of European ancestry, to investigate potential causal associations between increased fasting insulin adjusted for BMI (FI), erythrocytosis and its non-glycemic impact on HbA1c. We investigated the association between Triglyceride-glucose index (TGI), a surrogate measure of IR/HI, and glycation gap (difference between measured HbA1c and predicted HbA1c derived from linear regression of fasting glucose) in people with normoglycemia and preT2D.

**Results:** Inverse variance weighted MR (IVWMR) suggests increased FI increases haemoglobin (b=0.54+/-0.09, p=2.7 × 10^-10^), red cell count (RCC, b=0.54+/-0.12, p=5.38×10^-6^) and reticulocyte (RETIC, b=0.70+/-0.15, p=2.18×10^-6^). Multivariable MR indicates increased FI does not impact HbA1c (b=0.23+/-0.16, p=0.162) but reduces HbA1c after adjustment for T2D (b=0.31+/-0.13, p=0.016). Increased haemoglobin (b=0.03+/-0.01, p=0.02), RCC (b=0.02+/-0.01, p=0.04) and RETIC (b=0.03+/-0.01, p=0.002) might modestly increase FI. Increased TGI associates with decreased glycation gap, i.e. measured HbA1c was lower than expected based on fasting glucose, (b=-0.09±0.009, p<0.0001) in people with preT2D but not in normoglycemia (b=0.02±0.007, p<0.0001).

**Conclusions:** MR suggests increased FI increases erythrocytosis and might potentially decrease HbA1c by non-glycemic effects. Increased TGI, a surrogate measure of increased FI, associates with lower-than-expected HbA1 in people with preT2D. These findings merit confirmatory studies to evaluate its clinical significance.

## Introduction

The type 2 diabetes (T2D) pandemic is a major public health challenge which now affects more than 420 million people worldwide (1,2). Insulin resistance (IR) and hyperinsulinemia (HI) are early abnormalities in the pathogenesis of prediabetes (preT2D) and T2D (3). Close surveillance and timely intervention in people with IR/HI and preT2D can potentially prevent T2D and remit/improve glycemia in those who do develop T2D (4–6).

Increasingly, haemoglobin A1c (HbA1c) has replaced fasting glucose and/or the 75 g oral glucose tolerance test to diagnose preT2D, T2D and T2D remission. HbA1c is also used to set glycemic targets for people with diabetes (7–9). Advantages to using HbA1c compared to fasting glucose, include convenience and use of an assay that is standardized, stable, reproducible with limited intraindividual variability (1,10). It provides an average measure of glycemia in the preceding 2 to 3 months (1). However, altered red cell lifespan and erythrocytosis, which is not routinely assessed, can affect HbA1c measurement by non-glycemic pathways, which has implications in patients with red cell disorders and haemoglobinopathies (1,11). In people without T2D, including those with preT2D, non-glycemic parameters are a major predictor of HbA1c: higher haemoglobin associates with lower HbA1c (12,13). Observational studies have also shown an association between IR/HI and increased haemoglobin and red cell count (14–16), but whether this association is causal is not established, nor is it known whether this impacts HbA1c measurement through non-glycemic pathways.

Mendelian randomization (MR) can be used to infer potential causal associations between an exposure and an outcome by assessing the effects of genetic variants robustly associated with the exposure in one population, on the outcome of interest in a separate cohort (2 sample MR) (17). We undertook bidirectional MR to investigate potential causal associations between fasting insulin adjusted for BMI (FI) and erythrocytosis (haemoglobin, red cell count and reticulocyte count) in people of European ancestry using summary statistics from the largest genome wide association studies (GWAS). We undertook multivariable MR to assess the non-glycemic effects of FI on HbA1c after adjusting for elevated fasting glucose (FG) and type 2 diabetes (T2D). We also explored the association between triglyceride-glucose index (18), a surrogate measure of IR/HI and glycation gap (difference between measured HbA1c and predicted HbA1c from fasted glucose measurement) in people with normoglycemia and preT2D.

## Methods

### Cohorts

Please see Supplementary File 1 for information outlining cohorts used for MR analyses (19–25).

### Primary MR analyses

For our primary analysis we undertook bidirectional inverse variance weighted (IVW) MR with FI as exposure (Supplementary File 3) and Hb, red cell count (RCC), reticulocyte count (RETIC) as outcomes. A p value of <0.05 was considered significant for primary and secondary analyses. We followed the recently published STROBE-MR reporting guidelines (Checklist in Supplementary File 2)(26). As we used publicly available summary statistics from GWAS, we did not seek institutional approval. Informed consent was obtained from the investigators from each participant in the original study.

### Secondary analyses

As we found a potential causal association between FI and erythrocytosis, we undertook univariable MR to investigate the association between FI and HbA1c followed by multivariable MR adjusted for FG, T2D and Hb.

MR assumptions: MR is based on three assumptions. First, the instrument is robustly associated with the exposure, therefore we only used SNPs that were genome-wide significantly associated for all the instruments (17). Second that the instrument does not influence the outcome via another pathway other than the outcome i.e. no horizontal pleiotropy (17). Finally, the instrument is not influenced by any confounders (17). For univariable MR, we used inverse weighted MR (IVWMR) and additional sensitivity analyses including MR-Egger, weighted median, weighted mode and leave-one-out analyses.

IVWMR was performed by undertaking meta-analysis of the individual Wald ratio for each SNP in the instrument. By permitting a non-zero intercept, MR-Egger relaxes the assumption of no horizontal pleiotropy and returns an unbiased causal estimate, in the case of horizontal pleiotropy, providing that the horizontal pleiotropic effects are not correlated with the SNP-exposure effects (InSIDE assumption) (17,27). The median effect of all SNPs in the instrument was used for analysis using weighted median MR, which permits SNPs with a greater effect on the association to be evaluated by weighting the contribution of each SNP by the inverse variance of its association with the outcome: this is robust even if only 50% of the SNPs satisfy all three MR assumptions (28). Finally, SNPs were clustered into groups based on similarity of causal effects for weighted mode MR, with the cluster with the largest number of SNPs deriving the causal effect estimate (29). Cochrane’s Q test was used to assess heterogeneity, while leave-one-out analyses were conducted to assess if any MR estimate was biased by a single SNP potentially with horizontal pleiotropic effect (17) and the F statistic was calculated to assess the strength of the instrument exposure (17,30,31).

Univariable MR was conducted using the “TwoSampleMR” package in R (R studio® v1.3.1073 and R® v4.0.3). Linkage disequilibrium (LD) pruning was used to select a proxy (r^2^>0.8) if a SNP was not directly matched from the 1000 Genomes project (Version 0.5.6, Released 2021-03-35). The “ggplot2” and “metaphor” packages in R were used to create plots. We undertook inverse variance weighted multivariable MR (IVW Multivariable MR) to assess the effect of FI on HbA1c after adjustment for FG and T2D as well as Hb (32). Multivariable MR was conducted using both the “TwoSampleMR”, “Multivariable MR” and “RMultivariable MR” packages in R (R studio® v1.3.1073 and R® v4.0.3) where the latter two packages assessed heterogeneity via Cochrane’s Q test and strength of the instrument via F statistics (30,32). Plots were; generated using “plotobject”.

#### Overlap between exposure and outcome cohorts

There is no reported overlap between the cohorts.

### Observational Study

We received institutional approval from UHN research ethics board for the observational study. As we analyzed anonymized data, we did not obtain consent from individual patients. We undertook further analyses in a cohort of patients without T2D (n=7600 of whom 1096, i.e. 14.4%, have pre-T2D), who attended University Health Network (UHN) outpatient clinics between 2006 and 2022. We excluded patients who had attended diabetes clinics in the prior 2 years, those with fasting glucose &#x2265; 7mmol/L, HbA1c &#x2265; 6.5%, age >65 years or <18 years or Hb outside the sex-specific normal range. We did not undertake analyses in patients with diabetes as we did not have access to their medical records and could not ascertain the type of diabetes or their medications (e.g. insulin and sodium glucose co-transporter 1 inhibitors) which can impact both glycemia and erythrocytosis (33,34)

Using R studio® v1.3.1073 and R® v4.0.3, predicted HbA1c was assessed based on regression analysis of fasting glucose adjusted for age and sex. Glycation gap was calculated as the difference between measured HbA1c and predicted HbA1c. We assessed the association between triglyceride-glucose index(18), a surrogate measure of IR/HI, and the glycation gap. Triglyceride-glucose index (18) was calculated as ln[fasting triglyceride (mg/dL) × fasting plasma glucose (mg/dL)/2]. Correction factors of 88.57 and 18 were used to convert triglycerides to mg/L and fasting glucose to mg/dL, respectively.

The rms and lattice packages were used to fit a regression model and for estimation. The beta coefficient, standard error, y-intercept and p-value were analyzed in order to determine if there was an association between triglyceride-glucose index and glycation gap for all participants, those with pre-T2D (HbA1c 6-6.4% and fasting glucose 6-6.9 mmol/l) and those with normoglycemia (specified as HbA1c < 6% and fasting glucose <6 mmol/l). A p-value of < 0.05 was specified as being significant. The R^2^ and adjusted R^2^ were also analyzed to determine the fit of the model. An ANOVA table further analyzed if there was a linear relationship present.

Several diagnostic plots were created to test the presence of linearity and evaluate the fit of our model. A Normal Q-Q plot was created to test if the data had a normal distribution. If the data points fell onto a reasonably straight line, this would indicate a well fit model. Using the xyplot function, two plots were then created, both examining the residuals (residuals versus fitted values and residuals versus triglyceride-glucose index). If the plots produced a straight line, this would indicate a linear relationship. If the line was curved, this would indicate nonlinearity and splines would be required to analyze the cubic model.

Finally, the model was assessed for overfitting via validation. A set of random numbers was generated and the 0.632 Bootstrap method was used (35). The 0.632 Bootstrap method was chosen as to reduce bias by using correction factors. The R^2^ and mean squared error (MSE) were analyzed. An overfit model would produce a significantly different R^2^ and a higher MSE. Further, an optimism greater than 0.1 would suggest overfitting, as well as a slope with shrinkage. A decrease in g-index would also be suggestive of overfitting. It should be noted that MSE and g-index may be difficult to interpret as they vary based on sample size and range of data. The same analyses were undertaken for triglyceride-glucose index on Hb.

## Results

### 1. Primary analyses

#### Univariable MR Analyses of FI and erythrocytosis (Hb, RCC, RETIC)

Univariable inverse variance weighted MR suggests increased FI increases Hb (b=0.54+/-0.09, p=2.7×10^-10^, RCC (b=0.54+/-0.012, p=5.38×10^-6^) and RETIC (b=0.70+/-0.15, p=2.18×10^-6^), with concordant results with MR-Egger, weighted median, weighted mode and simple mode MR analyses. (Table 1, Figure 1-2).

**Table 1:**
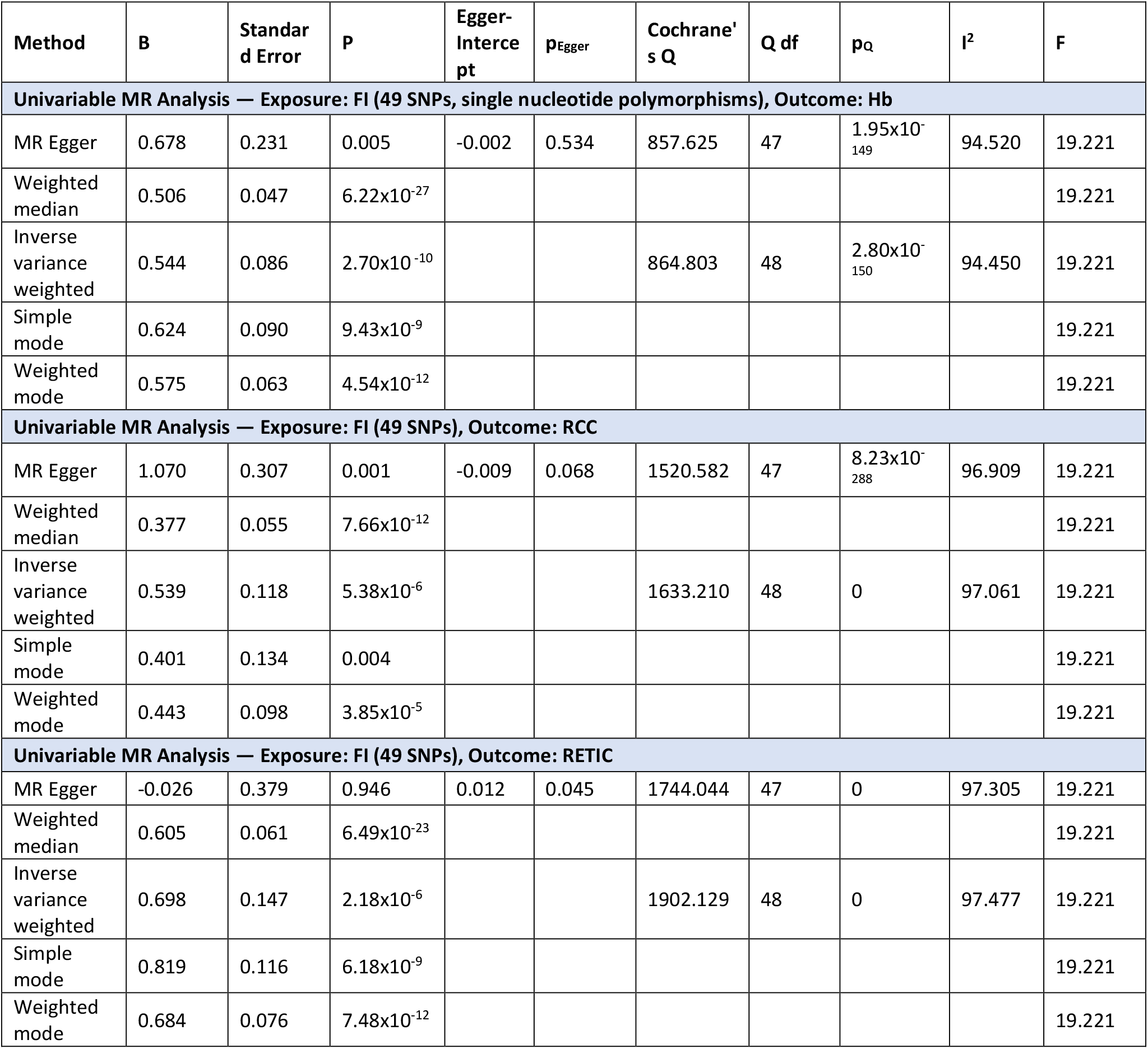
Univariable MR analyses of fasting insulin (FI) as exposure and haemoglobin (Hb), red cell count (RCC) and reticulocyte count (RETIC) as outcomes.

**Table 2:**
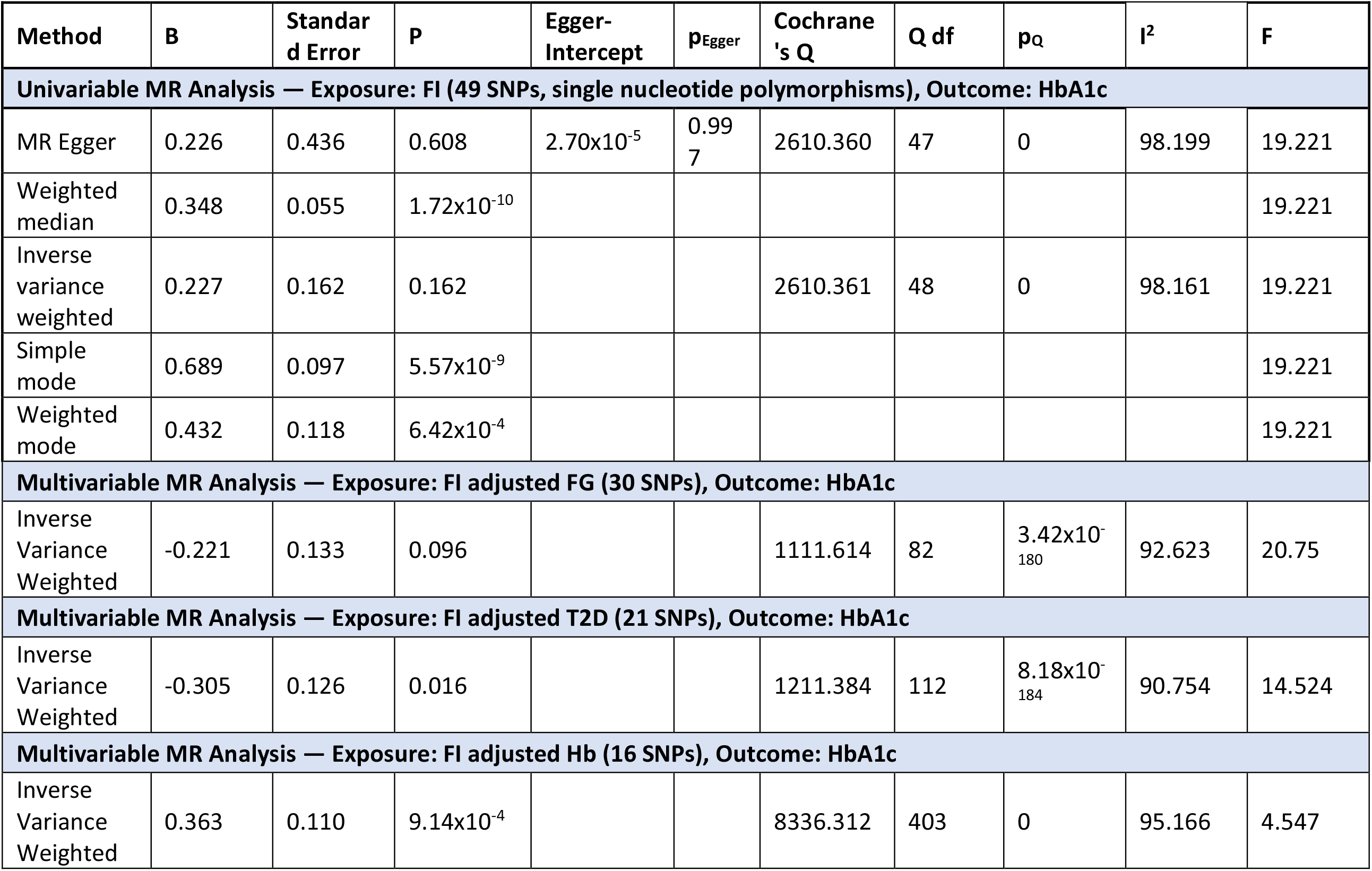
Univariable MR analyses of fasting insulin (FI) and glycated hemoglobin (HbA1c) as outcome followed by multivariable MR adjusted for elevated fasting glucose (FG), type 2 diabetes (T2D) and increased haemoglobin (Hb)

**Figure 1.** Univariable Mendelian Randomization (MR) Analysis — Exposure: fasting insulin (FI), Outcome: haemoglobin (Hb)— (A) Scatter plot showing the single nucleotide polymorphisms (SNPs) associated with FI against SNPs associated with Hb (vertical and horizontal black lines around points show 95% confidence intervals (CI) for five different Mendelian Randomization (MR) association tests (B) Funnel plot of the effect size against the inverse of the standard error for each SNP

**Figure 2.**
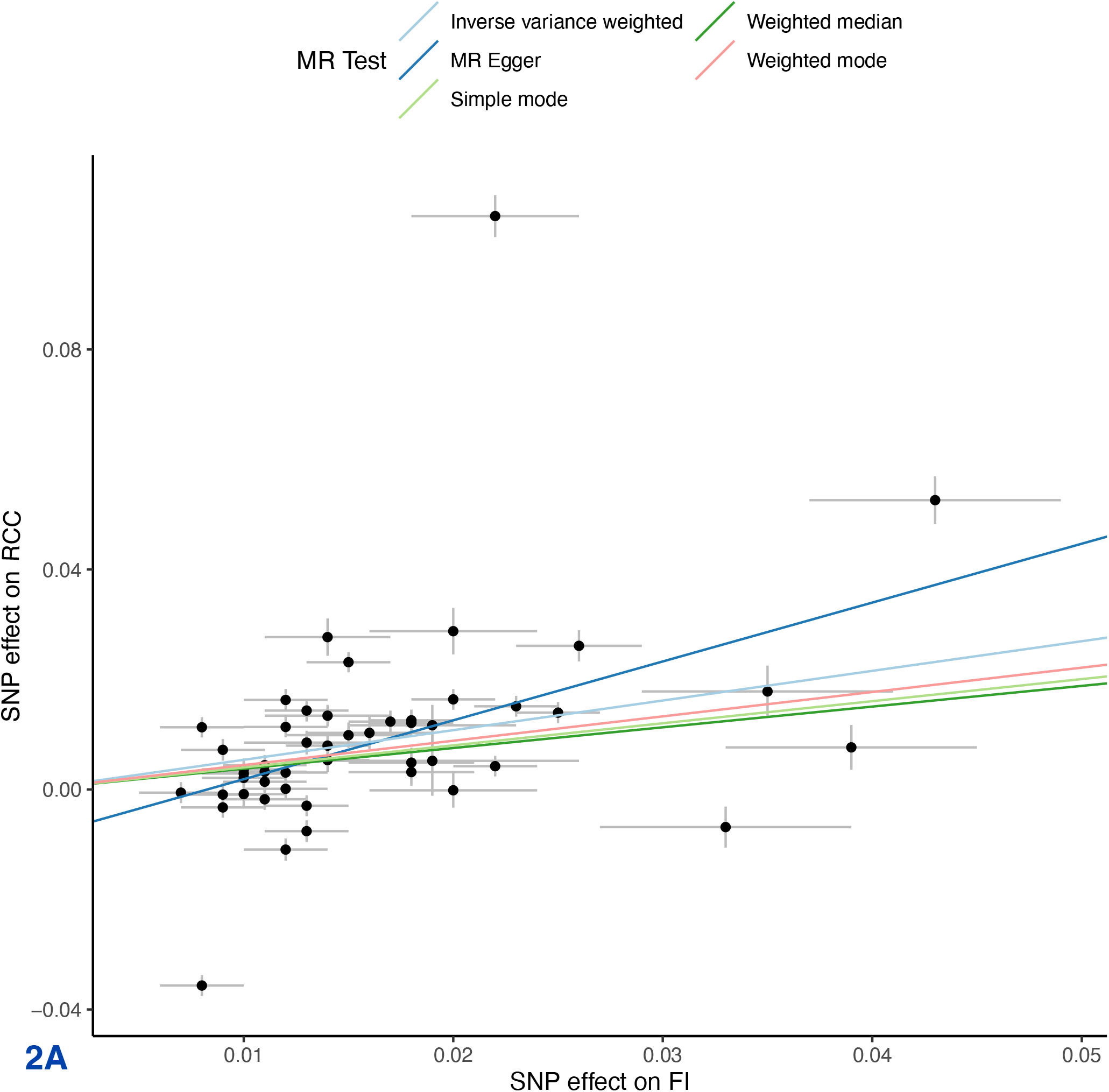
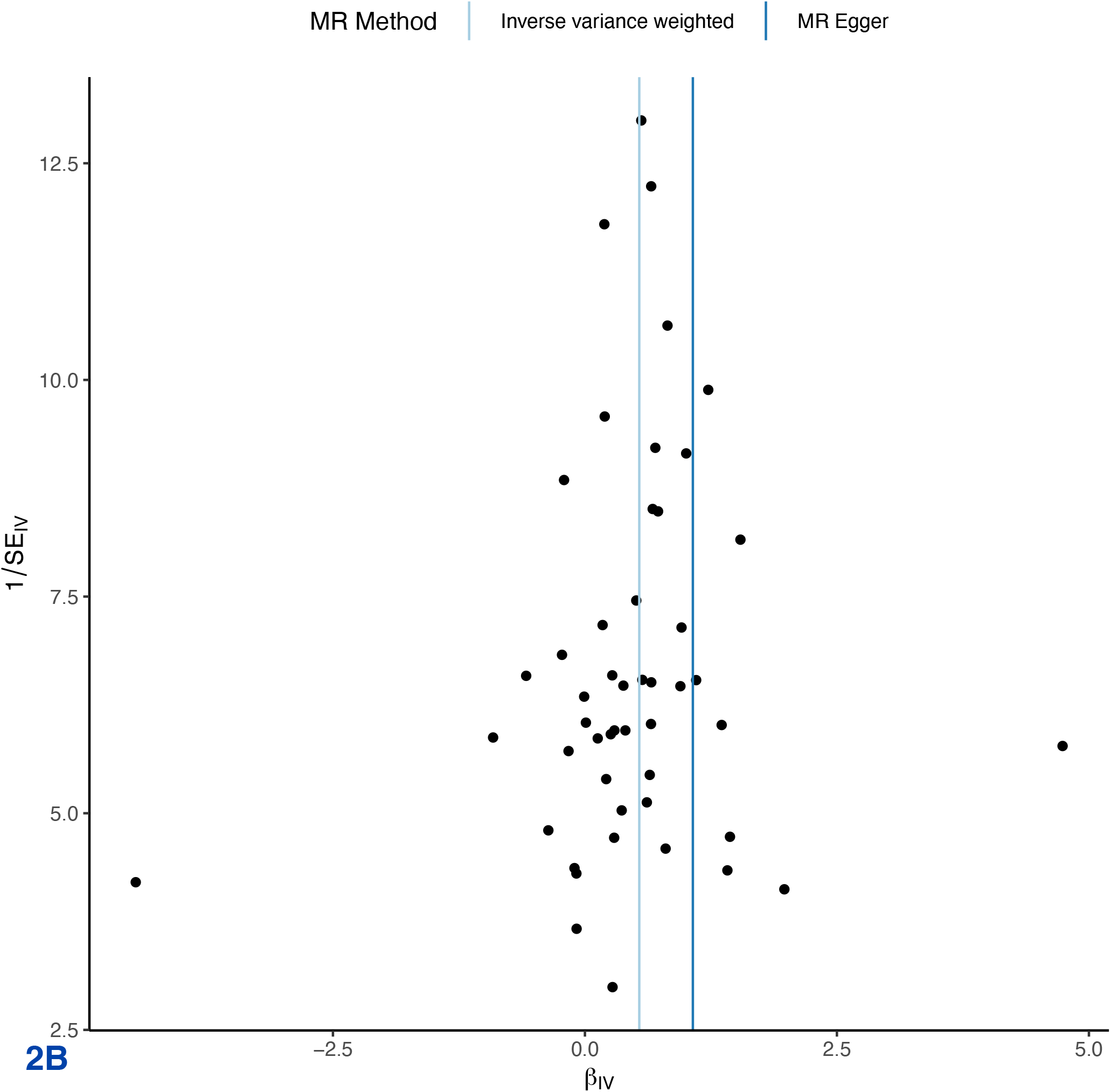
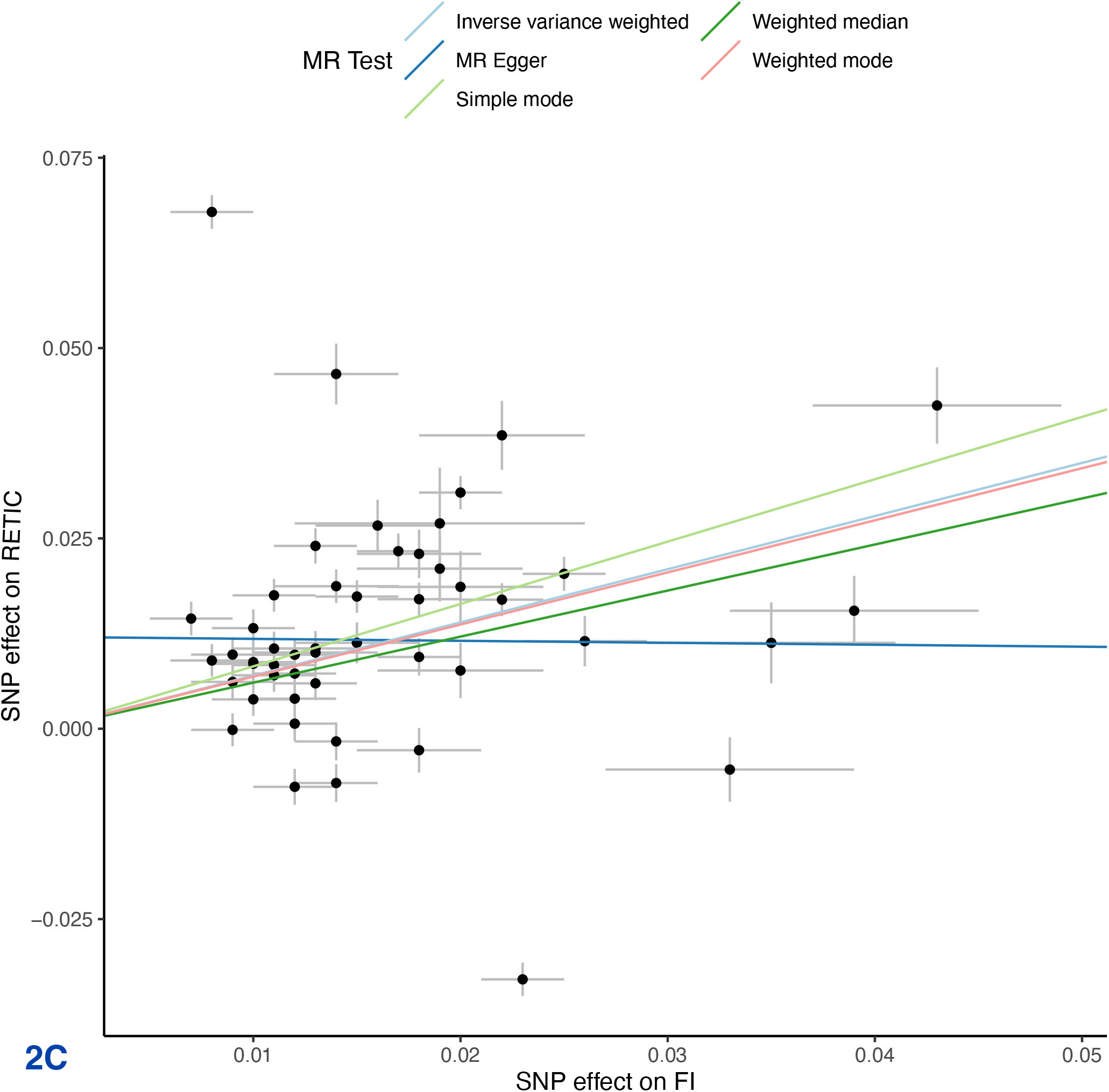
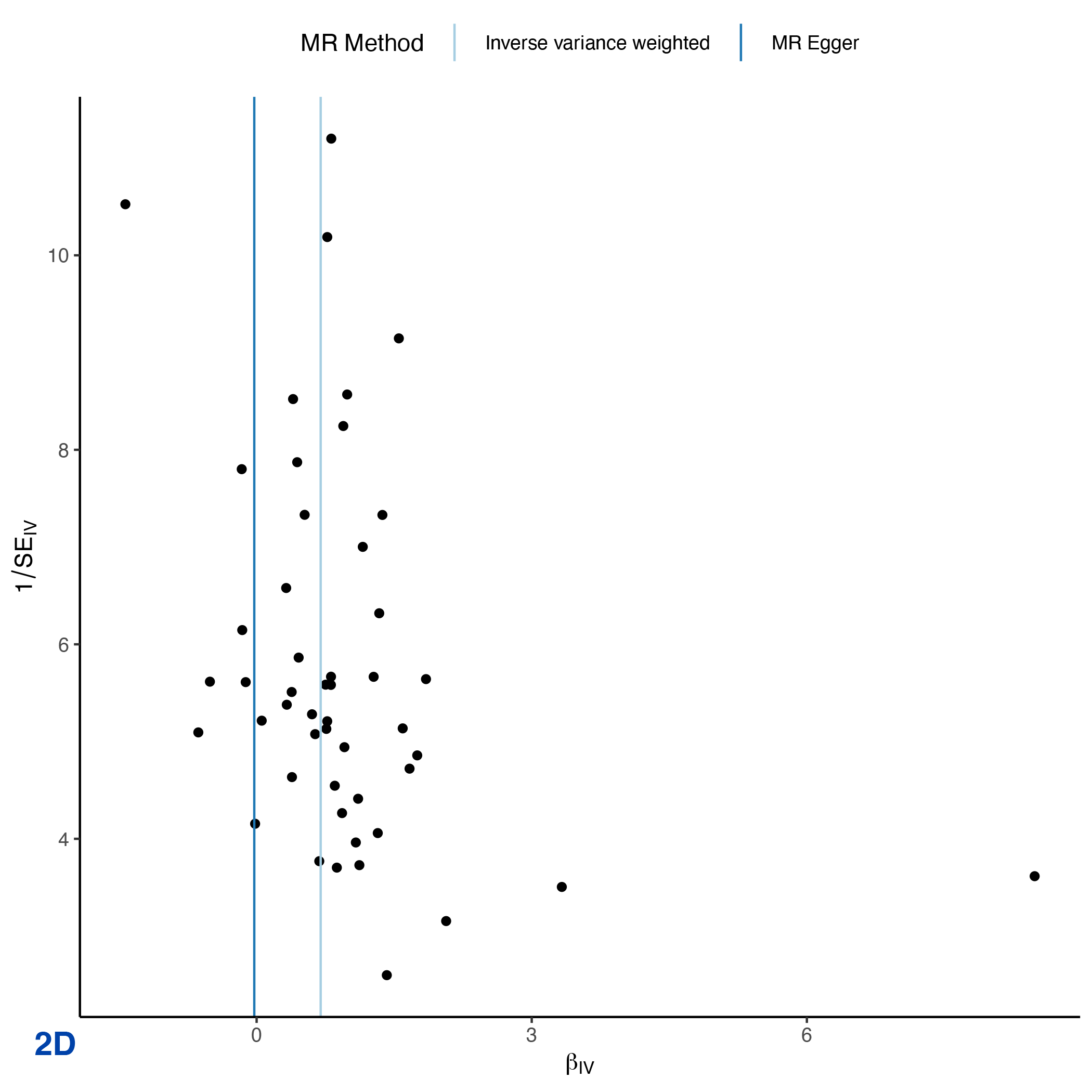
Univariable Mendelian Randomization (MR) Analysis — Exposure: fasting insulin (FI), Outcome: red cell count (RCC) and reticulocyte count (RETIC)— (A) Scatter plot showing the single nucleotide polymorphisms (SNPs) associated with FI against SNPs associated with RCC (vertical and horizontal black lines around points show 95% confidence intervals (CI) for five different Mendelian Randomization (MR) association tests (B) Funnel plot of the effect size against the inverse of the standard error for each SNP for FI against RCC (C) Scatter plot showing the single nucleotide polymorphisms (SNPs) associated with FI against SNPs associated with RETIC (vertical and horizontal black lines around points show 95% confidence intervals (CI) for five different Mendelian Randomization (MR) association tests (D) Funnel plot of the effect size against the inverse of the standard error for each SNP for FI against RETIC (

Inverse variance weighted MR suggests increased Hb (b=0.03+/-0.01, p=0.02), RCC (b=0.02+/- 0.01, p=0.04) and RETIC (b=0.03+/-0.01, p=0.002) might modestly increase FI, but MR-Egger, weighted median, weighted mode and simple mode MR analyses did not find evidence for potential causal association (Supplementary File 4)

### 2. Secondary analyses

#### Univariable and Multivariable MR Analyses of FI as exposure (adjusted for FG, T2D and Hb) and HbA1c as outcome

Univariable inverse variance weighted MR suggests that increased FI does not significantly increase HbA1c (b=0.23+/-0.16, p=0.16). Multivariable MR suggests FI decreases HbA1c after adjusting for T2D (b=-0.30+/-0.13, p=0.02. After adjusting for Hb (b=0.36, p=9.14×10^-4^), FI increases HbA1c, but the F-statistic of <10 precludes definitive conclusion. There was no significant effect of FI on HbA1c after adjusting for FG alone (b=-0.221+/-0.13, p=0.096) (Table 3, Figure 3, Supplementary File 5).

**Figure 3.**
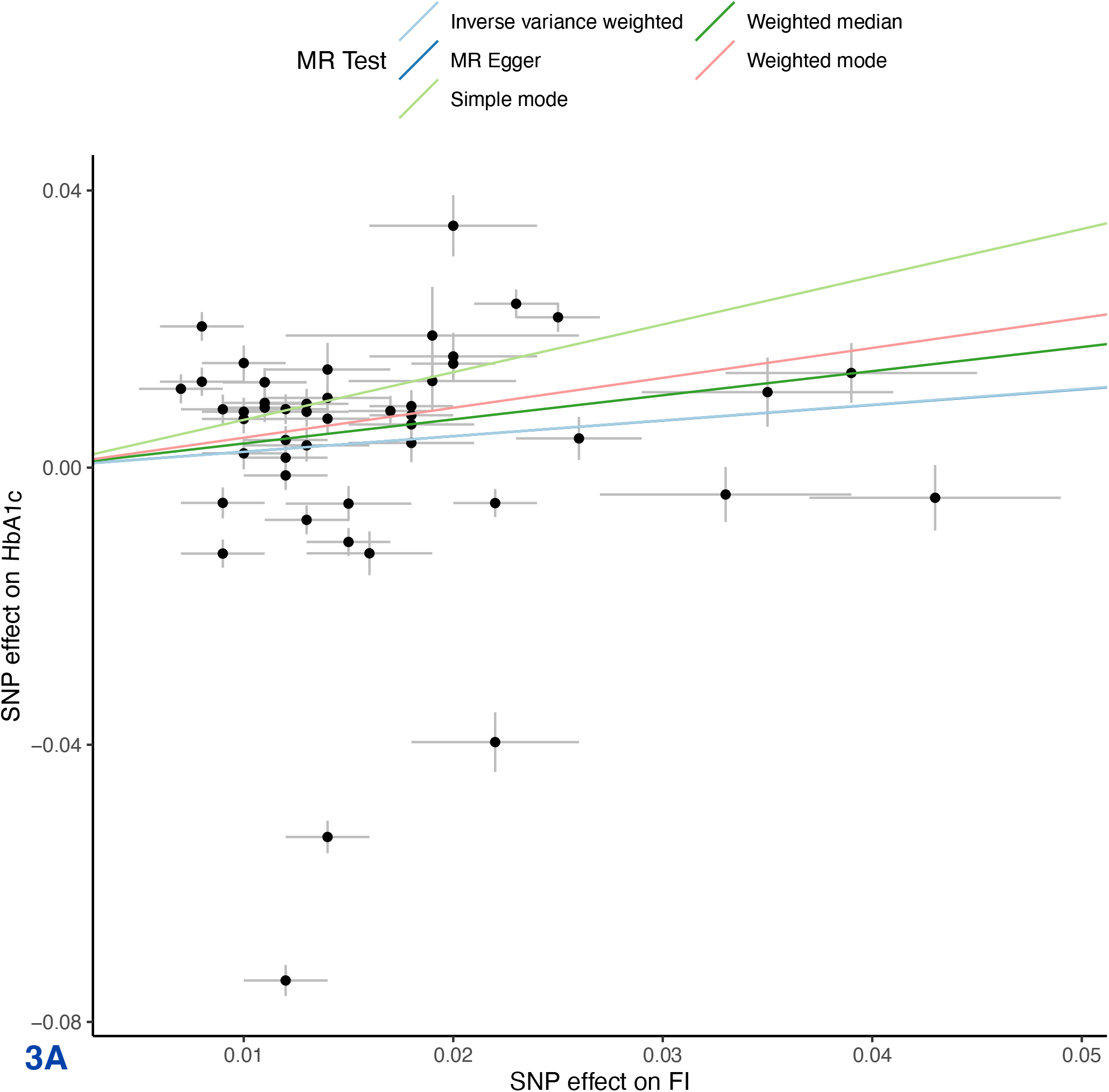
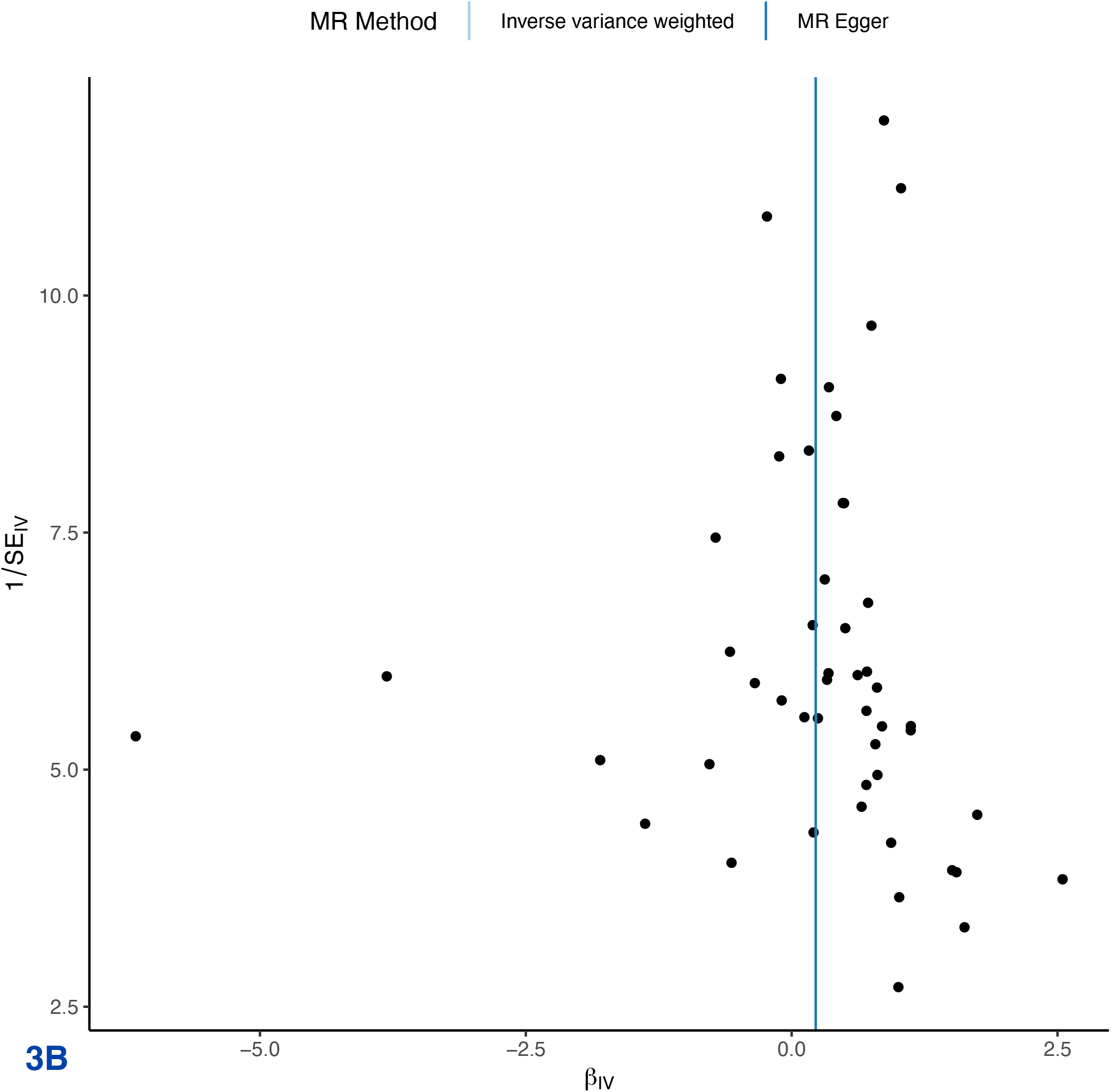
Univariable Mendelian Randomization (MR) Analysis — Exposure: fasting insulin (FI), Outcome: HbA1c— (B) Scatter plot showing the single nucleotide polymorphisms (SNPs) associated with FI against SNPs associated with HbA1c (vertical and horizontal black lines around points show 95% confidence intervals (CI) for five different Mendelian Randomization (MR) association tests (C) Funnel plot of the effect size against the inverse of the standard error for each SNP

### 3. Exploratory Analyses

#### MR Analyses exploring association between Hb and HbA1c

MR suggests a bidirectional relationship between Hb and HbA1c. Univariable inverse variance weighted MR suggests increased Hb decreases HbA1c (b=-0.105, p=1.17×10^-13^) concordant with MR-Egger, weighted median and mode but not simple mode analyses (Supplementary File 6). Reverse inverse variance weighted MR suggests increased HbA1c decreases Hb (b=-0.867, p=6.02×10^-7^) concordant with MR-Egger, weighted median and simple mode but not weighted mode analyses (Supplementary File 6).

#### Observational Study

Descriptive statistics regarding the 7600 participants can be found in Supplemental File 7. Consistent with the MR analyses, increased TGI was associated with increased Hb (b=1.88±0.19, p<0.001) (Figure 4a). Linear regression analysis yielded this equation for predicted HbA1c derived from fasting glucose and adjusted for age and sex: 4.163+0.172*(fasting glucose). In the cohort overall (the majority of whom had normoglycemia), (b=0.073±0.008, p<0.0001) and in people with normoglycemia (b=0.023+/-0.007, p<0.0001), increased triglyceride-glucose index was associated with an increase in glycation gap. However, among people with pre-T2D, increased triglyceride-glucose index was associated with a decreased glycation gap i.e.measured HbA1c was lower than that predicted by fasting glucose (b=-0.087+/-0.009, p<0.0001) (Figure 4b-d).

**Figure 4.**
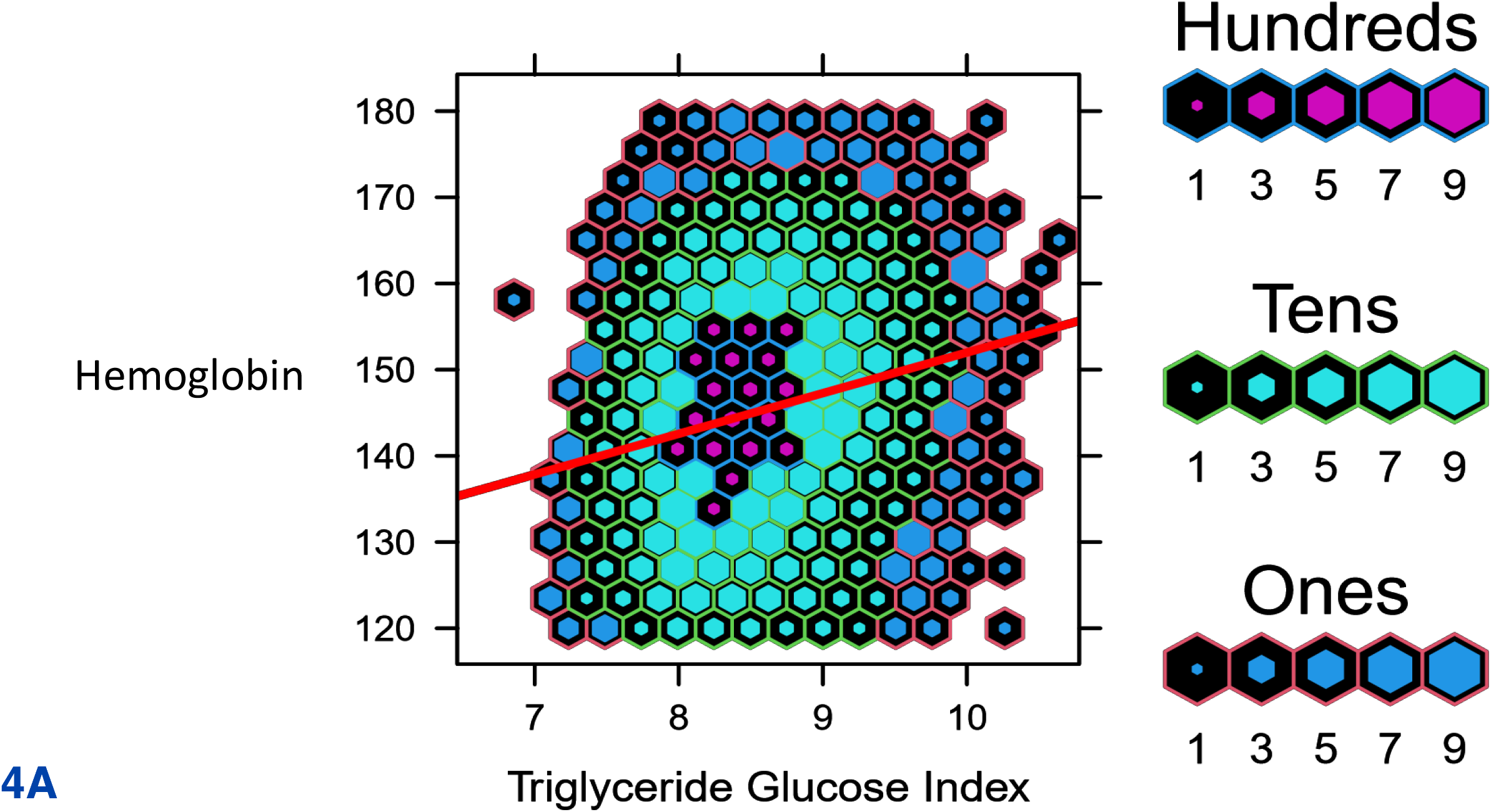
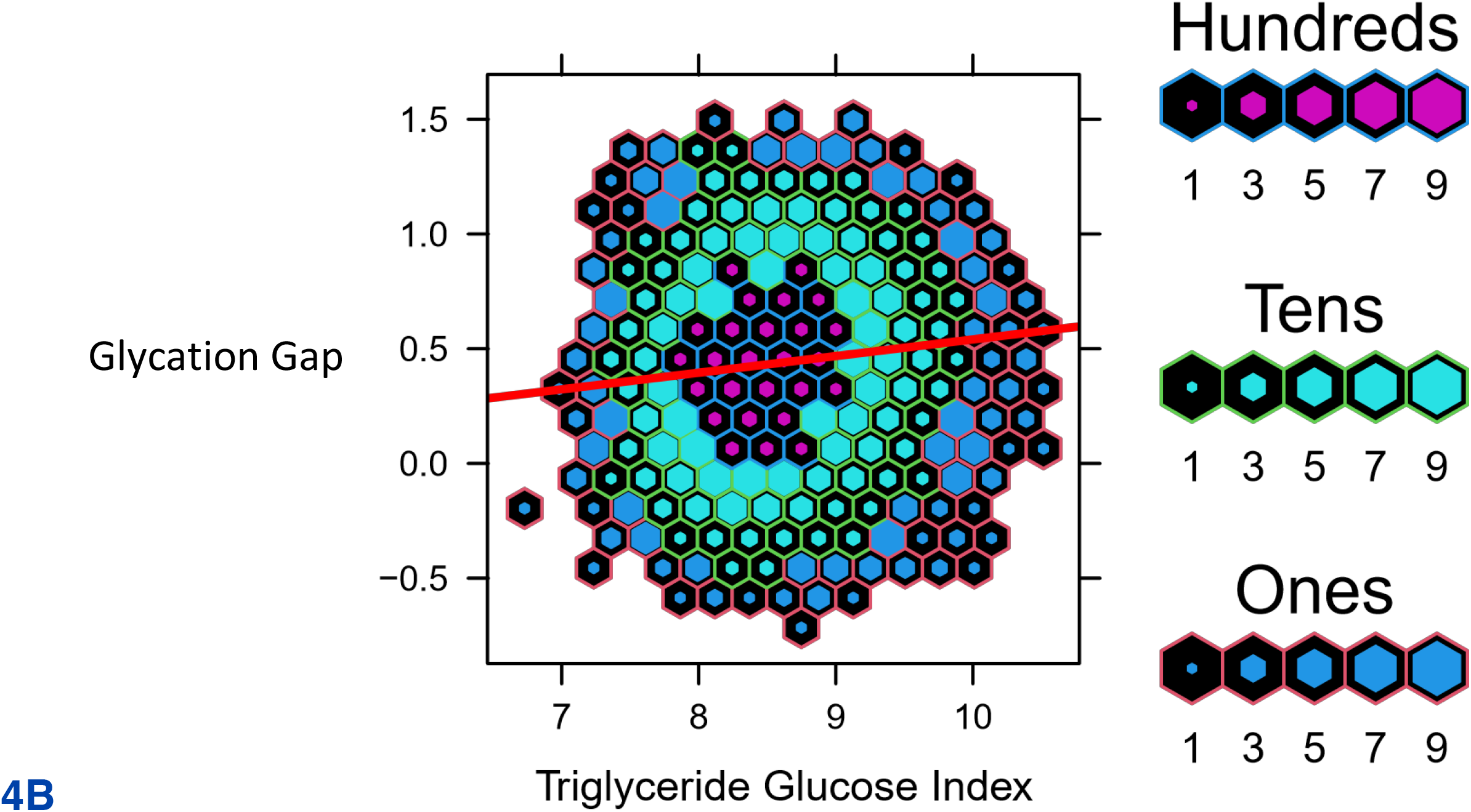
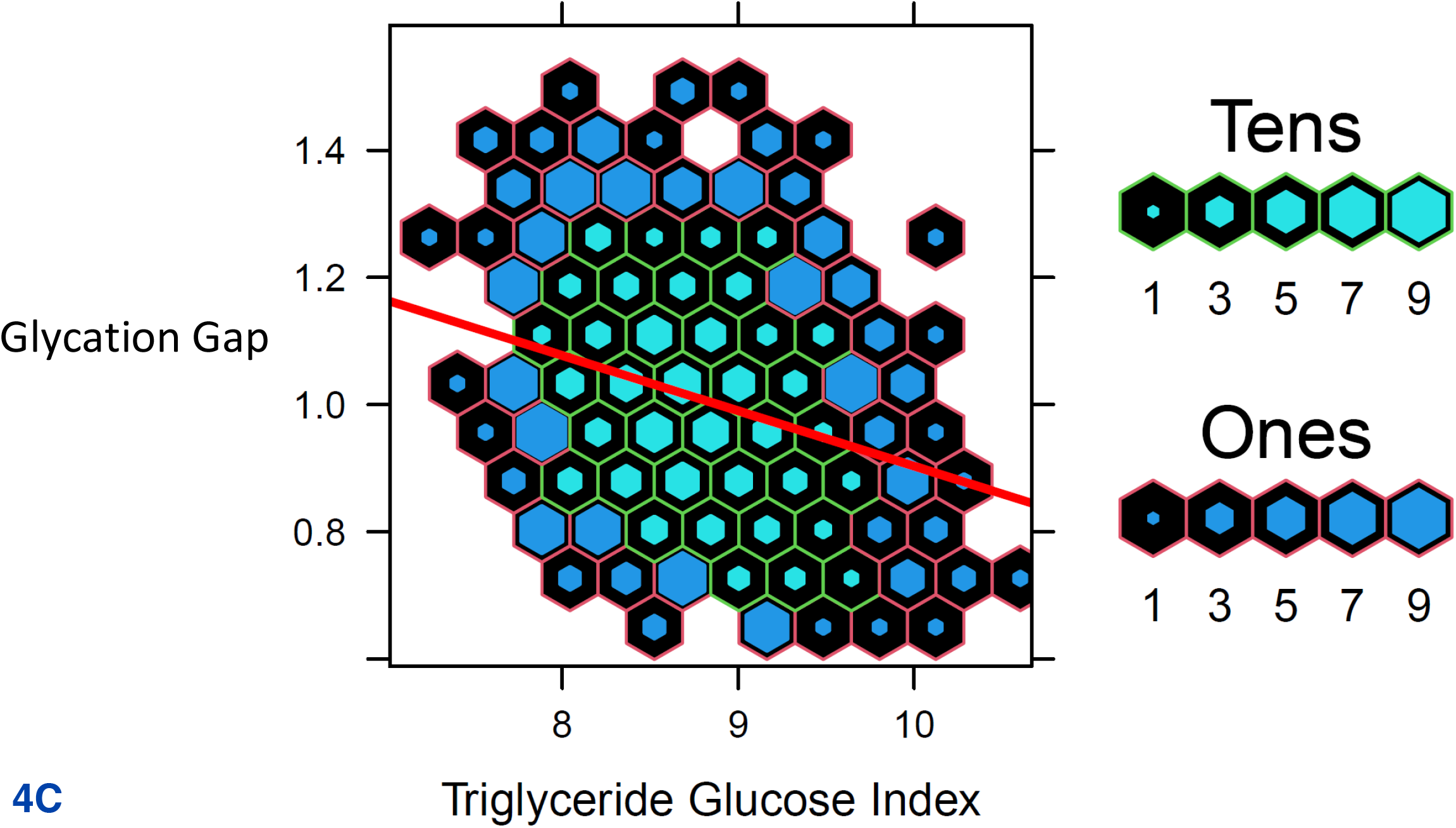
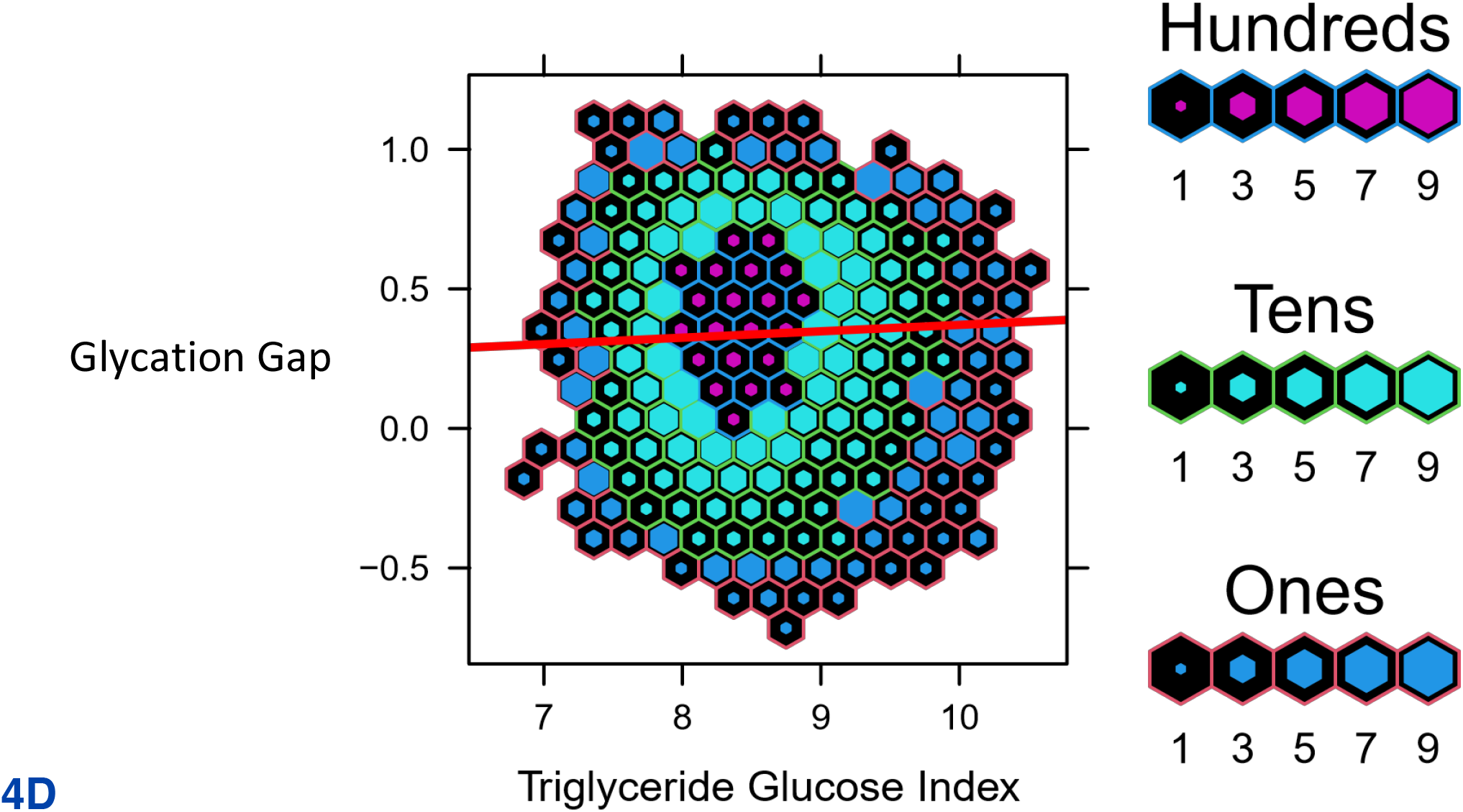
Data from UHN cohort. Hexbin plots represent number of study participants with the observed and calculated values, where the colour and size of each individual hexagon correlates to the number of participants with the corresponding values. The red line represents the regression line for each cohort. (A) Association between Triglyceride Glucose Index and Hemoglobin (B) Association between Triglyceride Glucose Index and Glycation Gap in all participants (C) Association between Triglyceride Glucose Index and Glycation Gap in with pre-T2D (D) Association between Triglyceride Glucose Index and Glycation Gap in those with normoglycemia.

For all analyses, the R^2^ adjusted and unadjusted were almost identical and the regression p-values were <0.005. Normal Q-Q plots for all data were suggestive of a normal distribution and good fit. Plots for residuals versus triglyceride-glucose index and residuals versus fitted values suggested linearity and that likely all relationships were accounted for in the model. Using the 0.632 Bootstrap method, validation was carried out and resulted in corrected R^2^ and corrected slopes that were relatively similar to the original values. The R optimism was found to be 0.0116, which was less than the cut-off of 0.1. The MSE increased by 1.57% and the g-index decreased by 3.97%. These results indicate that the model was not overfit.

## Discussion

Epidemiological data suggests that IR/HI is associated with increased erythrocytosis (14–16). Our MR analysis suggests that this association is causal. MR further suggests that increased FI after adjustment for T2D reduces HbA1c. MR also indicates a bidirectional inverse relationship between Hb and HbA1c. Collectively, these data suggest that increased IR/HI mediated erythrocytosis might potentially lower HbA1c by non-glycemic effects with the transition from normoglycemia to T2D. Our observational data was concordant with the MR analyses. shows Increased triglyceride-glucose index, a surrogate measure of IR/HI associated with both higher Hb and measured HbA1c which is lower than expected in people with preT2D, but not in those with normoglycemia. These findings await confirmation and assessment of clinical significance in well designed prospective studies across the glycemic spectrum from normoglycemia to preT2D and T2D.

Increased FI is a recognised manifestation of IR. Some features of IR/HI such as increased hepatic glucose production are likely a consequence of reduced insulin action, while others such as hepatic steatosis and dyslipidemia are likely due to increased insulin action via signaling pathways that are not perturbed in IR (36). *In vitro* studies suggest that insulin can increase erythrocytosis (37), suggesting that increased insulin action may underlie the increased in erythrocytosis with IR/HI. Further studies are needed to confirm these findings and explore underlying mechanisms.

The potential non-glycemic impacts of increased fasting insulin on HbA1c might lead to lower-than-expected HbA1c based on glycemia and thus have implications for people with IR/HI and preT2D and T2D. HbA1c is increasingly used to diagnose these conditions, in lieu of fasting glucose/oral glucose tolerance test measures, and to set glycemic targets for treatment (1,7–10). Interestingly, observational data indicate that ∼40% of people with T2D diagnosed based on more than one measure of elevated fasting glucose and/or post OGTT glucose, have HbA1c below the diabetes threshold (38). The potential non-glycemic effects of increased FI on HbA1c may also be particularly pertinent for weight loss induced T2D diabetes remission. HbA1c is the recommended glycemic parameter to define remission in a patient population with high prevalence of IR/HI (9).

The strengths of this study include MR analyses with the largest sample sizes in populations of European ancestry and likely minimal/no overlap between participants in the exposure and outcome cohorts. Our study has a number of limitations. The findings may not apply to other ethnic groups given that we used populations with European ancestry only. This may especially be a concern in populations with higher prevalence of hemoglobinopathies and red cell disorders (39–43). Additionally, analyses were not stratified by sex, which is a major determinant of body composition and IR/HI (44). For our observation data we did not have access to individual level data including medications and comorbidities. Due to these limitations, we also excluded patients with biochemical evidence of T2D as we could not reliably ascertain the type of diabetes and account for the potential impact of medications which might impact red cell parameters. We derived predicted HbA1c from fasting glucose and did not account for post-prandial readings which are a major limitation. Finally, we did not have measures of FI in the observational cohort and therefore used surrogate measures of IR/HI in our analyses.

In conclusion, our data suggests that increased FI may increase erythrocytosis and might potentially lower HbA1c independent of glycemia. As these findings might have implications for the diagnoses and management of preT2D and T2D, it merits well designed prospective confirmatory studies across the glycemic spectrum to confirm these findings and assess whether these effects are clinically relevant.

## Data Availability

All data produced in the present work are contained in the manuscript

## Funding

SD is funded by CIHR, Heart & Stroke Foundation of Canada and Banting & Best Diabetes Centre (DH Gales Family Charitable Foundation New Investigator Award and a Reuben & Helene Dennis Scholar in Diabetes Research).

## Contributions

SD, ADP, AN and RK designed the study. All authors analyzed the data. AN, RK and SD wrote the manuscript and all authors read and edited the manuscript. SD is the guarantor of this work.

## Conflicts of Interest

None

## Supplementary File 1 – Cohorts

Summary statistics from the largest published genome wide association study (GWAS) in people of European ancestry were used in MR analyses (Supplementary Table 1) (18–24). Informed consent and institutional approval were previously obtained by the individual cohort investigators.

**Table:** Cohort details (all participants were of European descent)

**Table: Cohort details (all participants were of European descent)**
| Trait | Population cohort | Mean Age | % female | Sample size | Cases | Controls | PMID |
| --- | --- | --- | --- | --- | --- | --- | --- |
| Fasting Insulin (FI) | MAGIC | 50.7 | 51.2 | 151,013 | N/A | N/A | 34059833 |
| Waist-to-hip ratio (WHR) | GIANT/UK Biobank | 55.5/56.9* | 54.0/54.2* | 694,649 | N/A | N/A | 30239722 |
| Type 2 Diabetes (T2D) | DIAGRAM/GERA/UK Biobank | 54.1/63.3/56.9* | 50.1/59.0/54.2* | 655,666 | 61,714 | 593,952 | 30054458 |
| Fasting Glucose (FG) | MAGIC | 50.9 | 47.7 | 133,010 | N/A | N/A | 22885924 |
| Haemoglobin (Hb) | UK Biobank | 56.7 | 54.9 | 563,946 | N/A | N/A | 32888493 |
| Red Cell Count (RCC) | UK Biobank | 56.7 | 54.9 | 545,203 | N/A | N/A | 32888493 |
| Reticulocytes (RETIC) *** | UK Biobank | 56.7 | 54.9 | 408,112 | N/A | N/A | 32888494 |
| HbA1c** | MAGIC | 52.3 | 57.9 | 146,806 | N/A | N/A | 34059833 |
| HbA1c**/** | UK Biobank | 56.7 | 54.9 | 389,889 | N/A | N/A | 34017140 |

\*Study-specific characteristics were not available for all UK Biobank data and was extrapolated from data available.
<sup>a</sup>Output from MRC IEU GWAS pipeline analysis using Pheasant derived variables from UK Biobank, version 2: https://doi.org/10.5523/bris.pnoat8cxo0u52p6ynfaekeigi
\*\* To minimize overlap, bidirectional MR analyses with FI was undertaken with HbA1c measure in the UK Biobank, but for WHR adjusted for BMI analyses HbA1c was assessed in MAGIC.
\*\*\* Estimated from available UK Biobank data (PMID 32888493) as data not available

## Supplementary File 2. STROBE-MR checklist of recommended items to address in reports of Mendelian randomization studies^1 2^

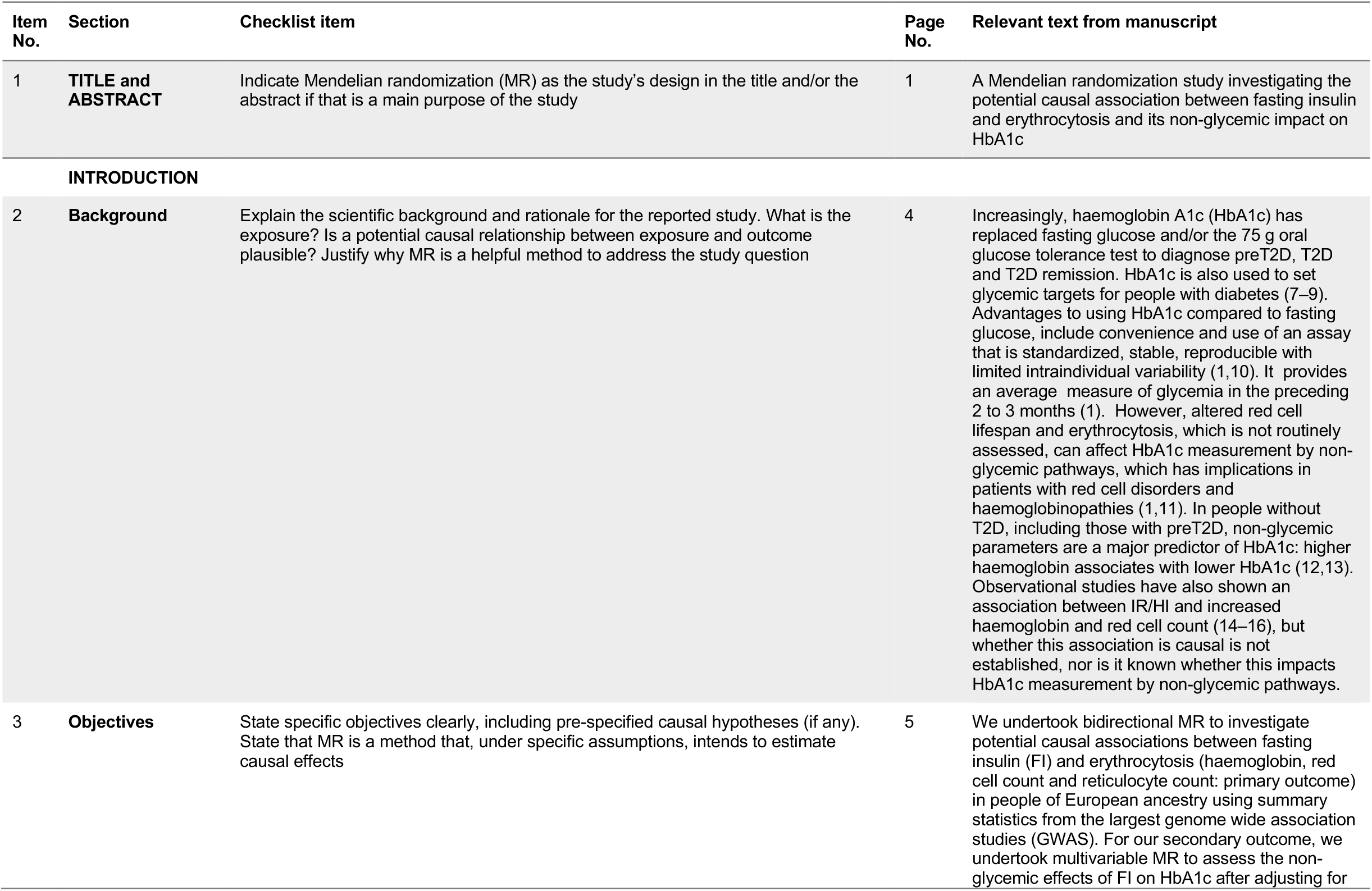
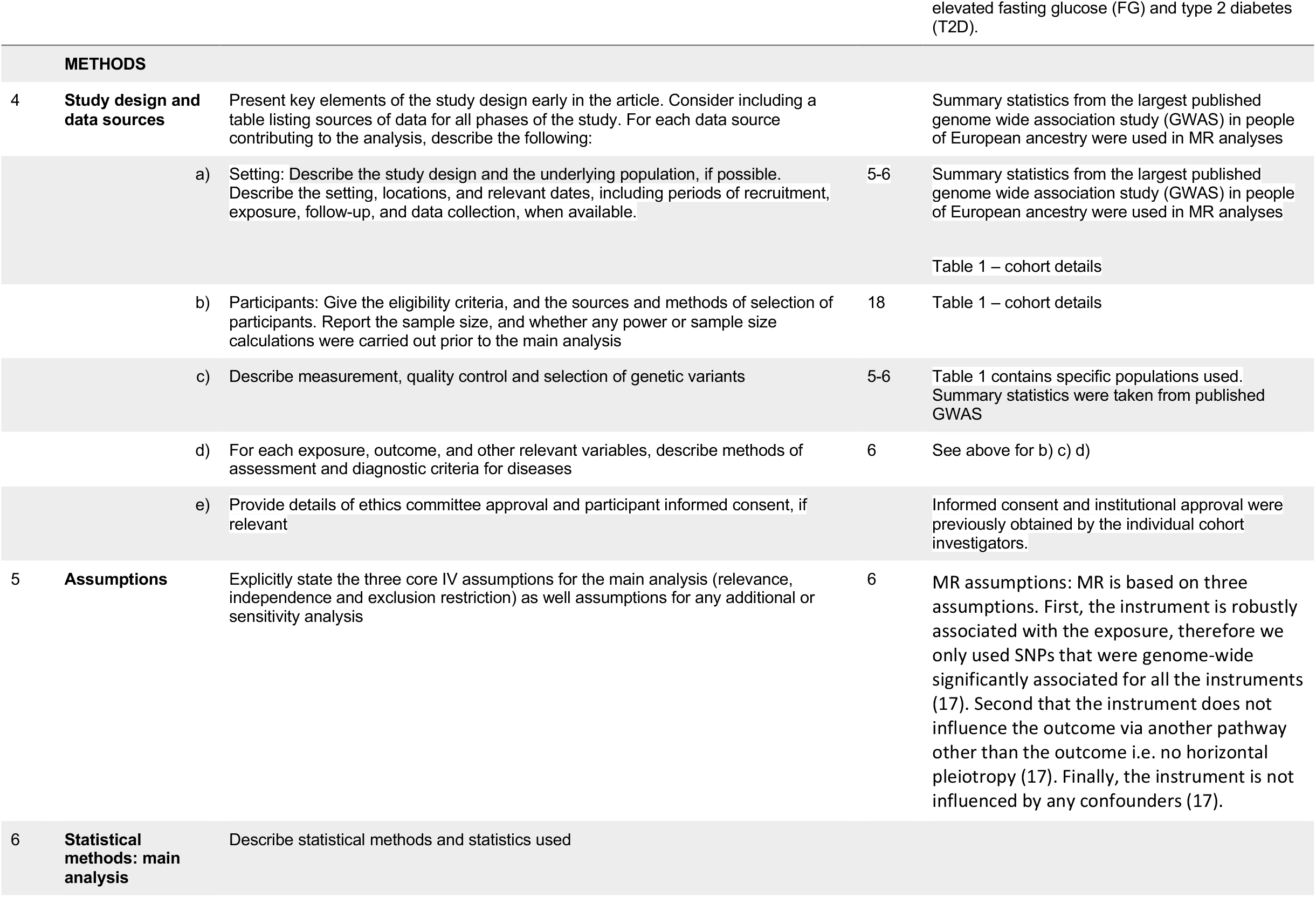
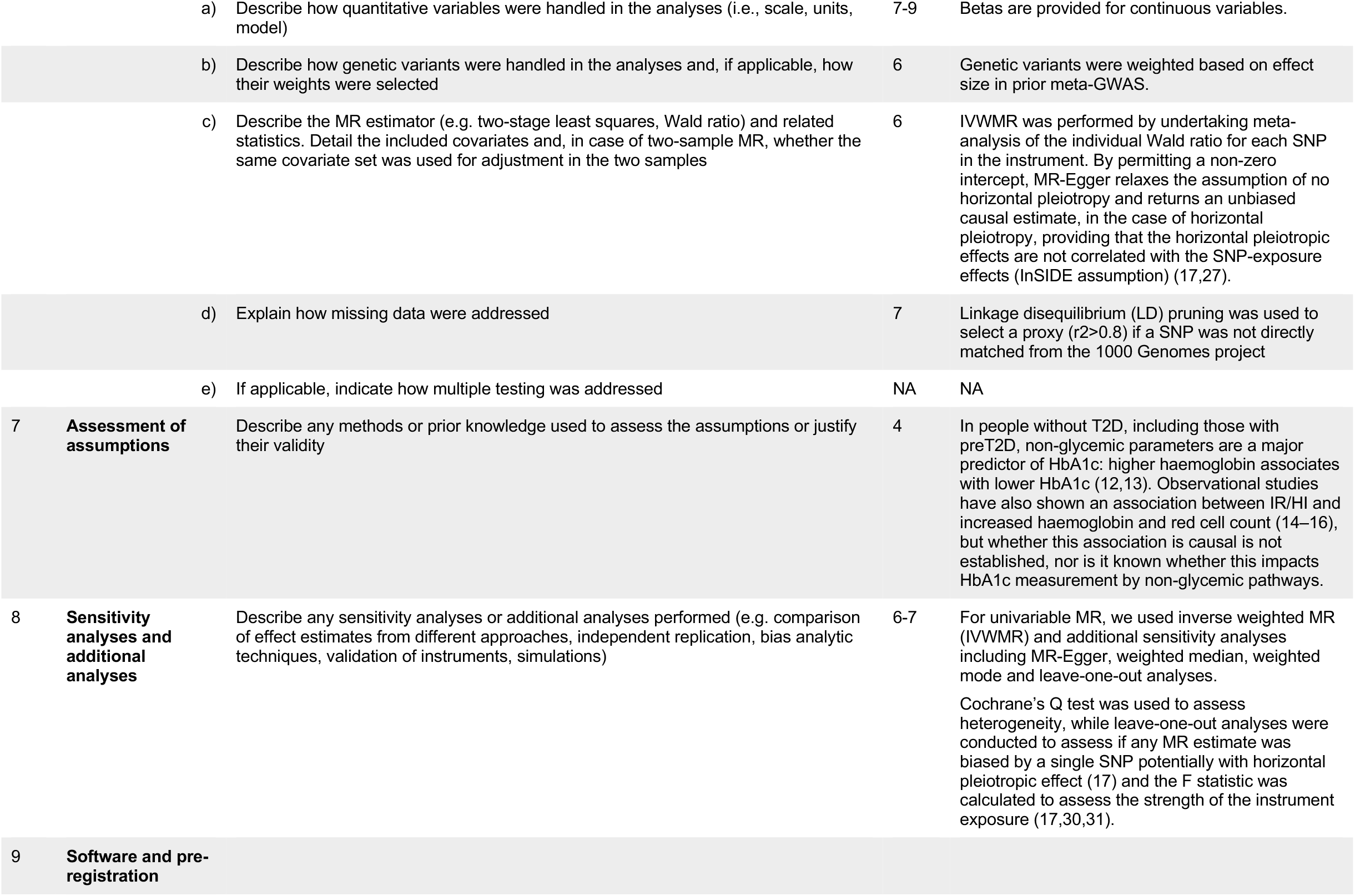
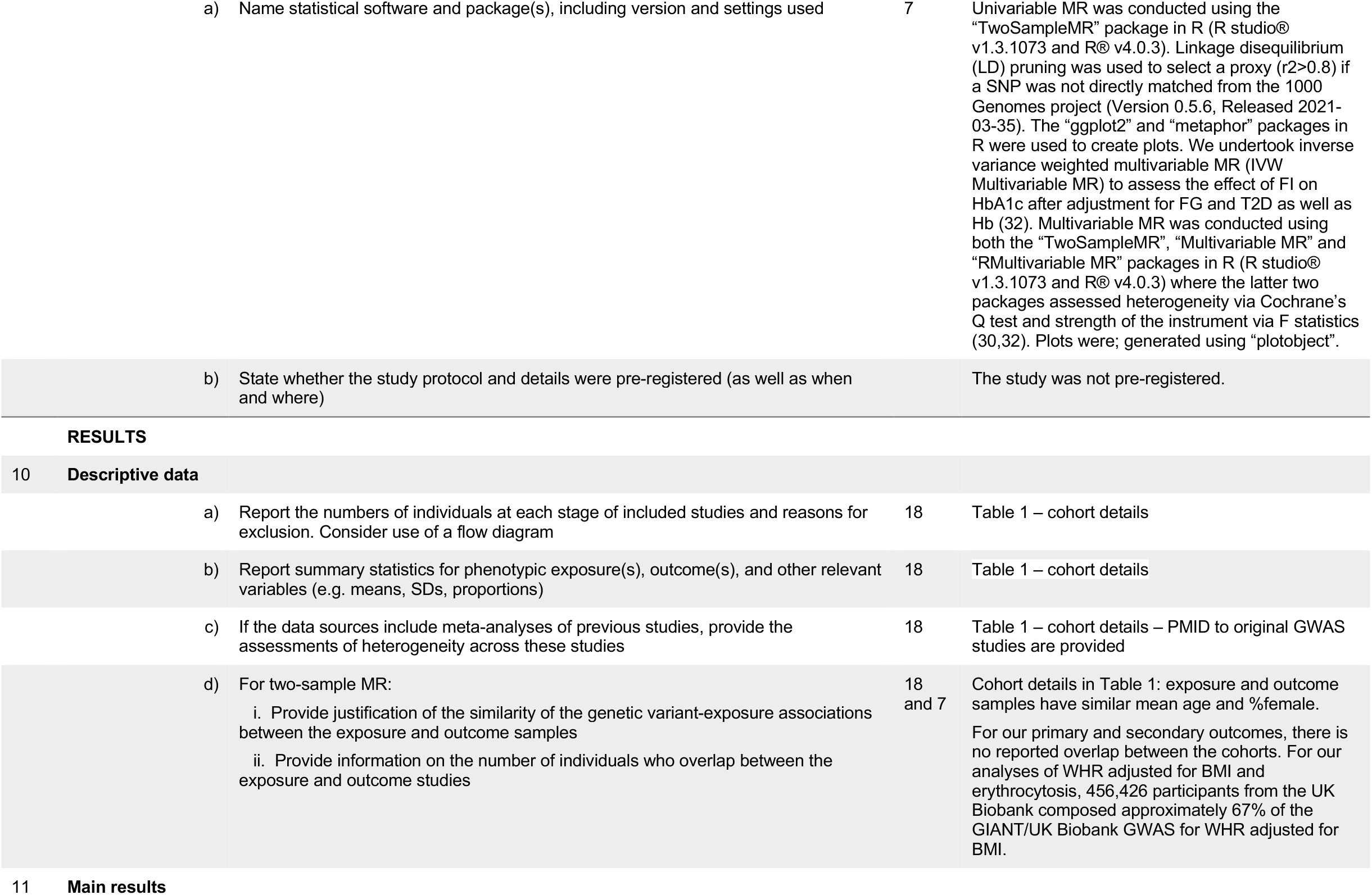
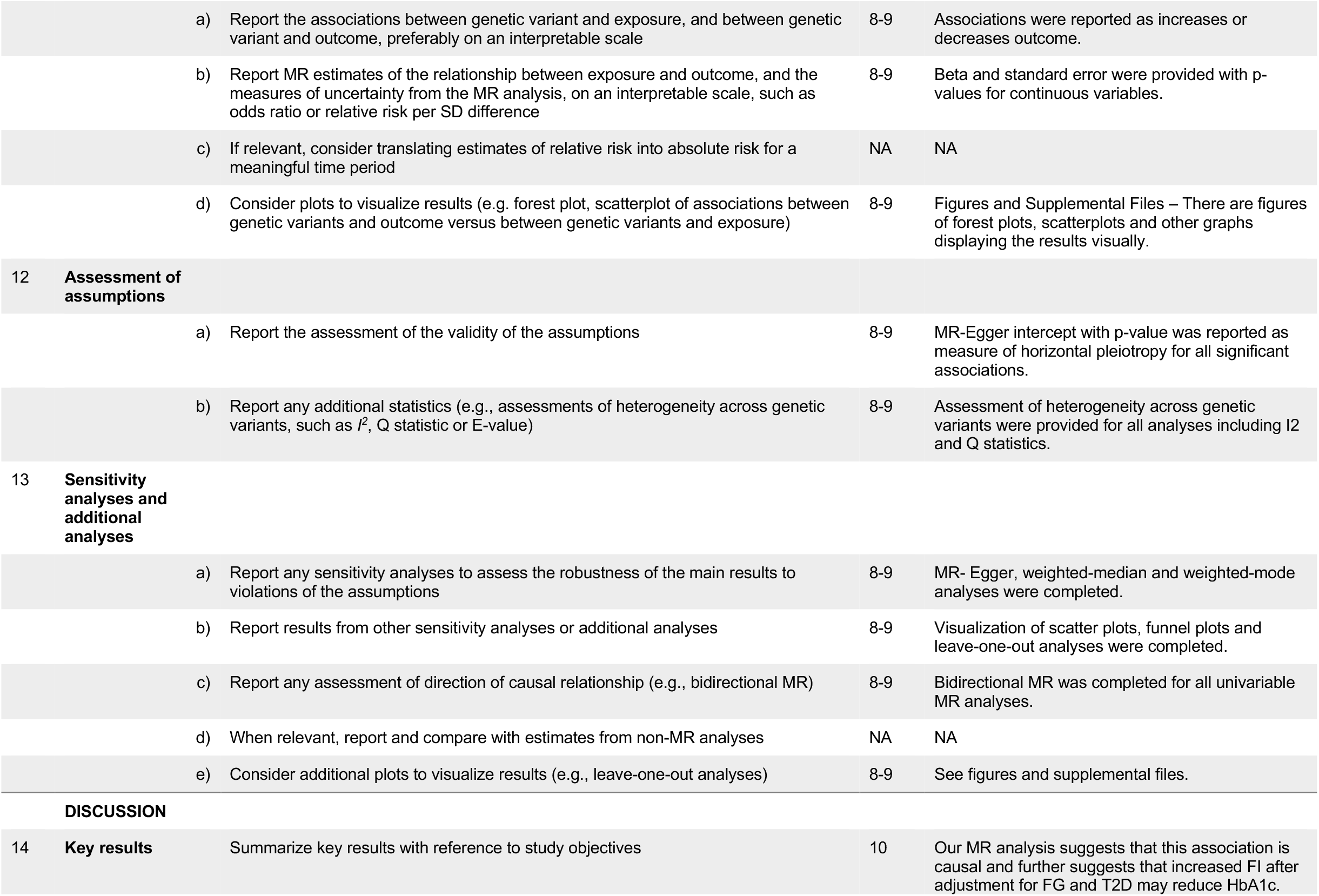
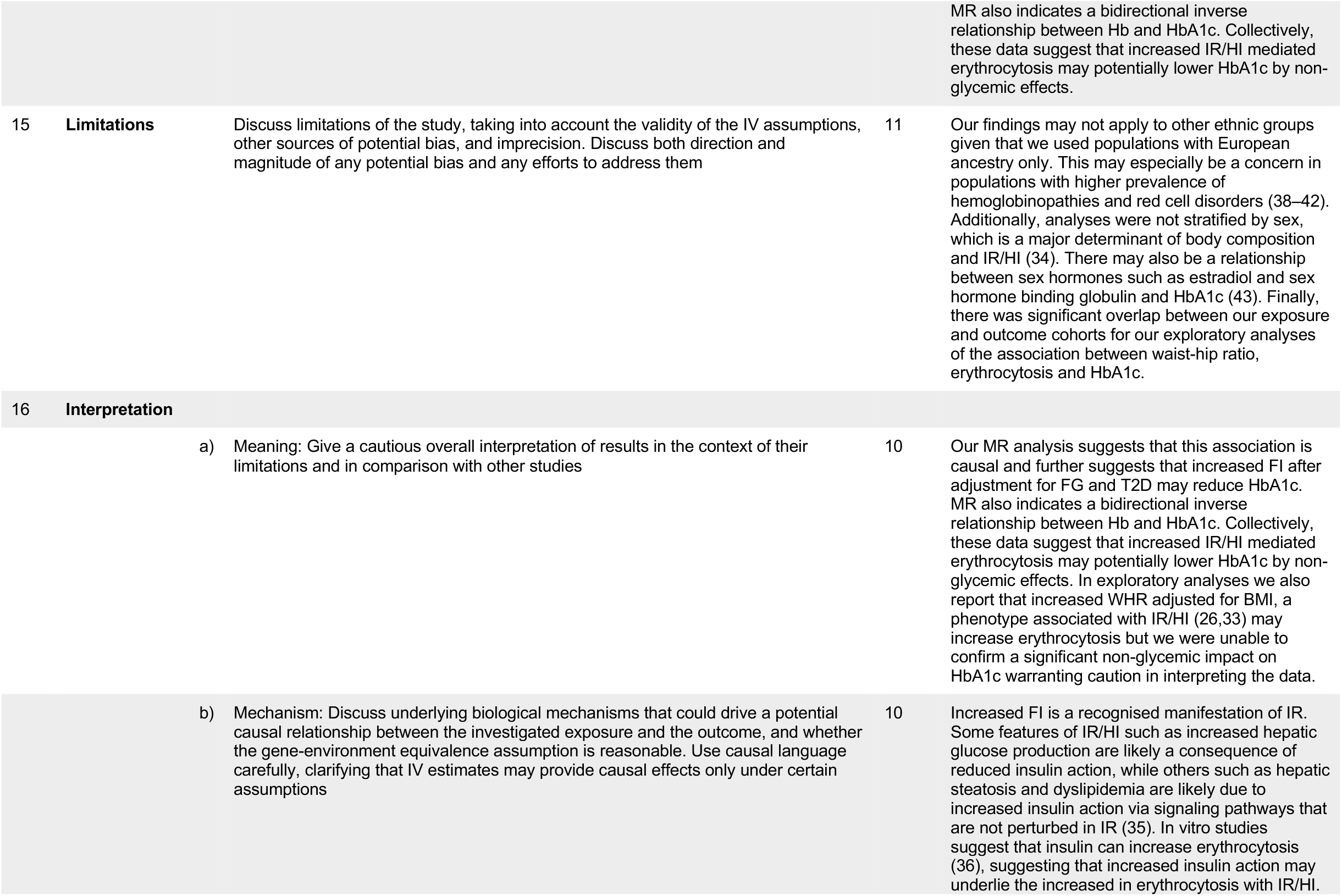
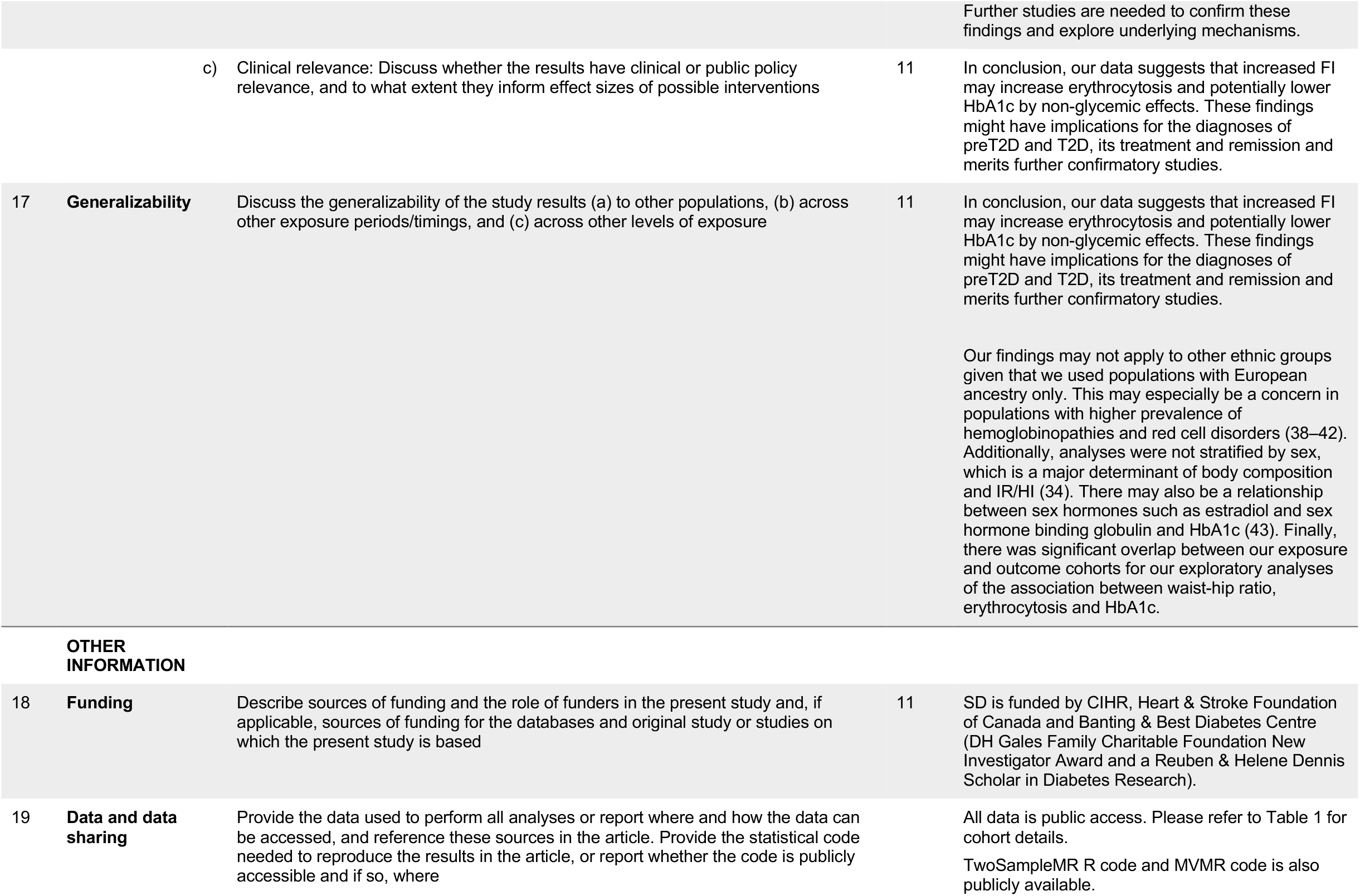
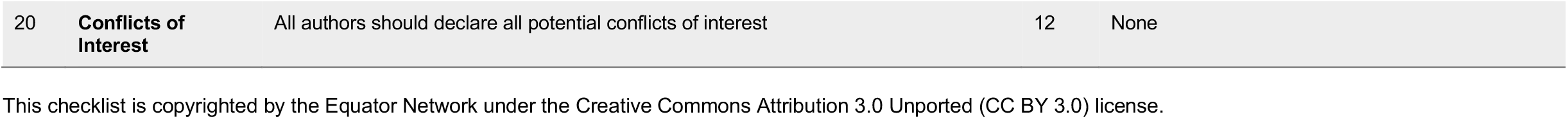

## Supplementary File 3. Fasting Insulin instrument

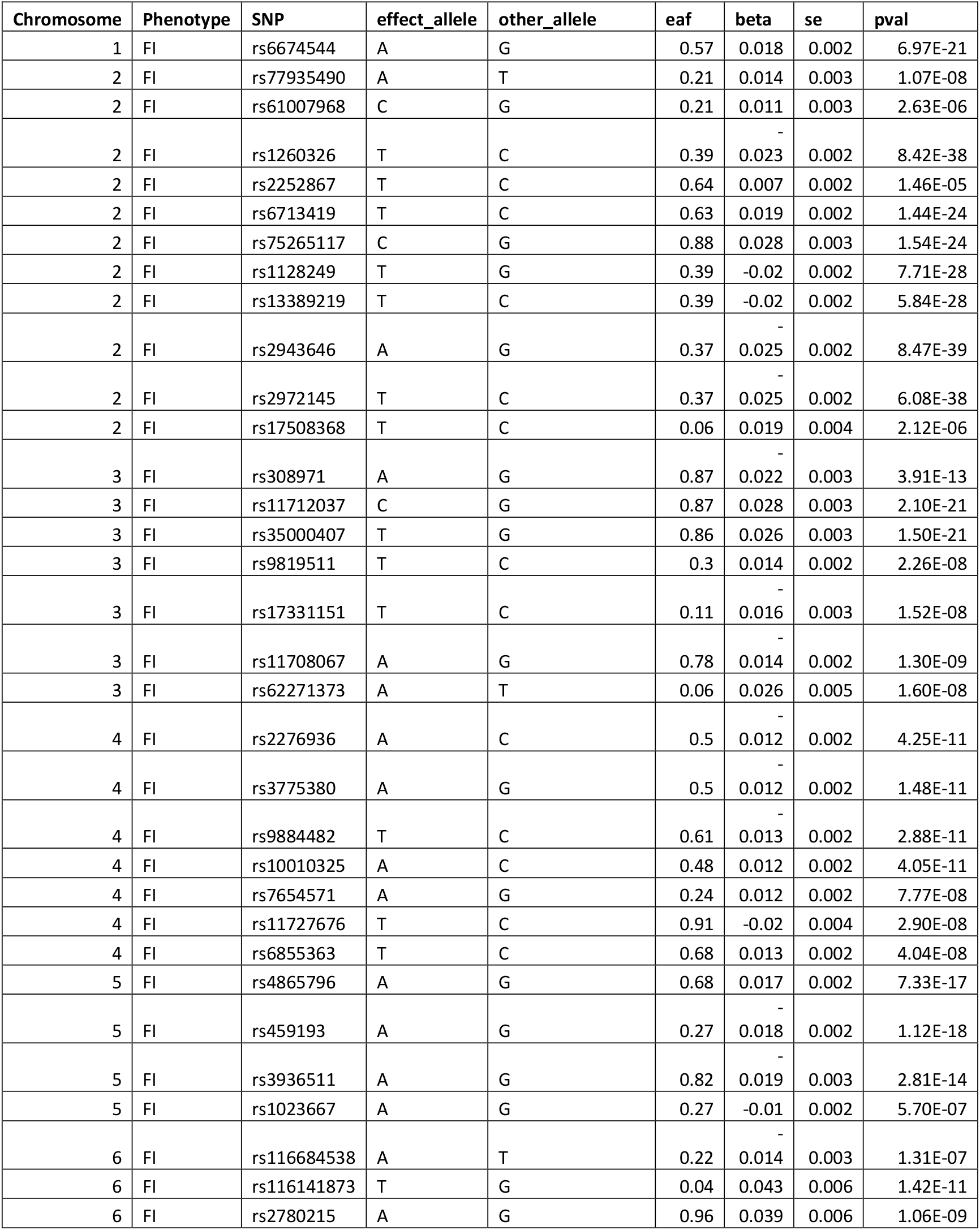
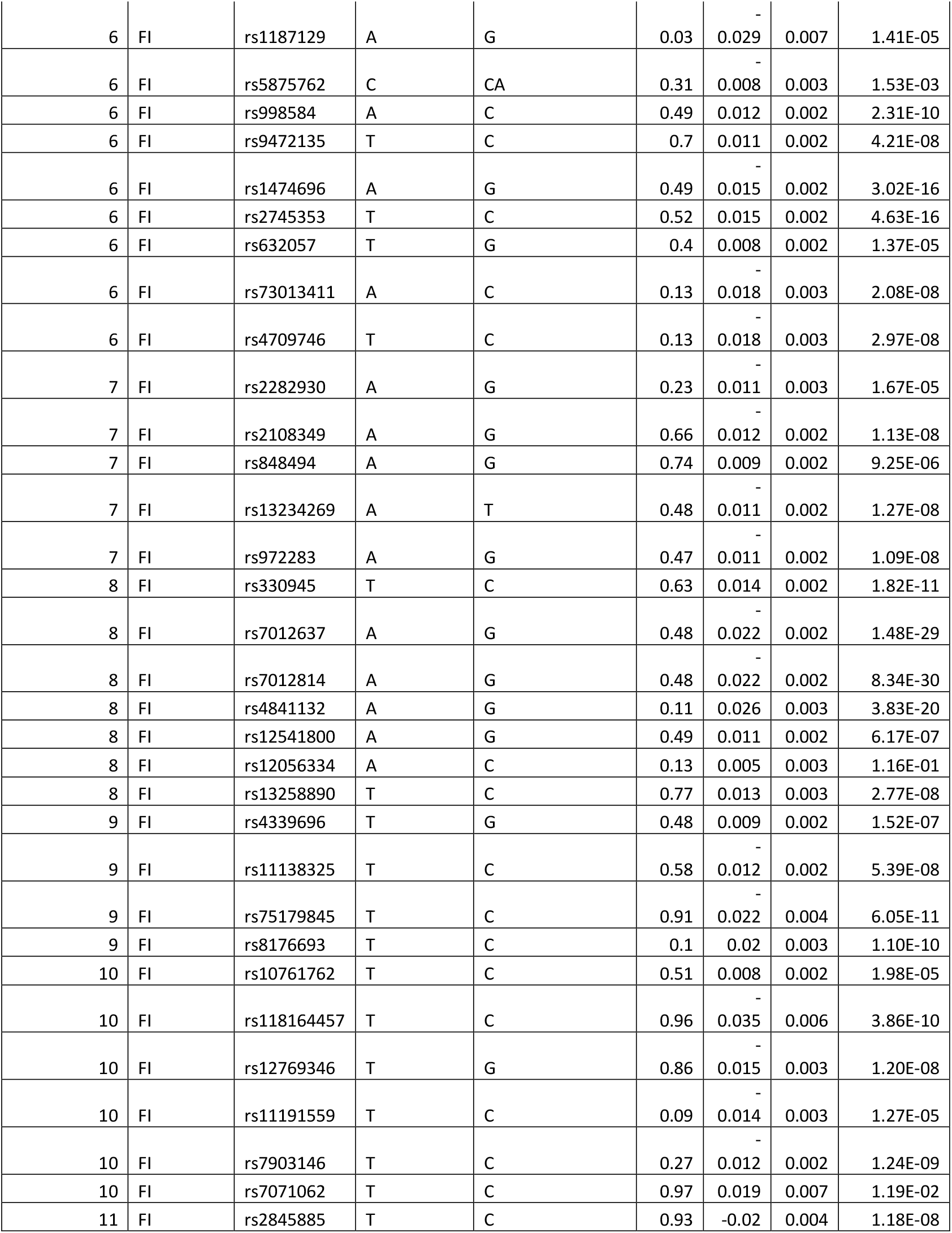
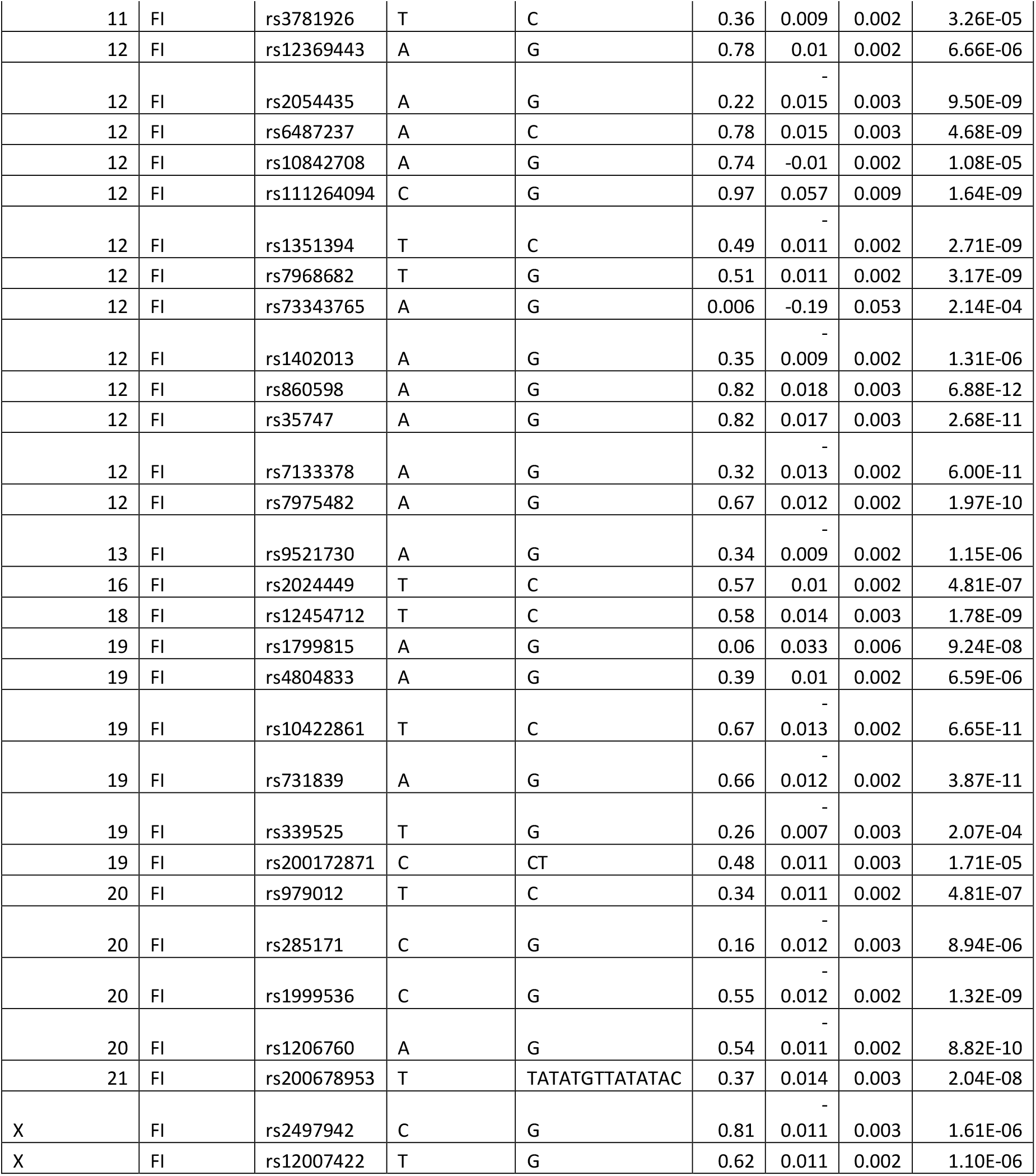

## Supplemental File 4: Reverse MR analyses. Exposure: Hb, RCC and RETIC. Outcome: Fasting insulin

| Method | B | Standard Error | P | Egger-Intercept | p<sub>Egger</sub> | Cochran's Q | Q df | p<sub>Q</sub> | I<sup>2</sup> | F |
| --- | --- | --- | --- | --- | --- | --- | --- | --- | --- | --- |
| **Univariable MR Analysis — Exposure: Hb (184 SNPs, single nucleotide polymorphisms), Outcome: FI** |  |  |  |  |  |  |  |  |  |  |
| MR Egger | -0.008 | 0.026 | 0.746 | 0.001 | 0.055 | 805.243 | 182 | 3.33x10<sup>-79</sup> | 77.398 | 95.738 |
| Weighted median | -0.021 | 0.013 | 0.098 |  |  |  |  |  |  | 95.738 |
| Inverse variance weighted | 0.033 | 0.014 | 0.017 |  |  | 821.763 | 183 | 1.13x10<sup>-81</sup> | 77.731 | 95.738 |
| Simple mode | -0.032 | 0.029 | 0.266 |  |  |  |  |  |  | 95.738 |
| Weighted mode | -0.020 | 0.012 | 0.102 |  |  |  |  |  |  | 95.738 |
| **Univariable MR Analysis — Exposure: RCC (214 SNPs), Outcome: FI** |  |  |  |  |  |  |  |  |  |  |
| MR Egger | -0.014 | 0.021 | 0.504 | 0.001 | 0.050 | 736.709 | 212 | 3.93x10<sup>-59</sup> | 71.223 | 147.934 |
| Weighted median | 0.000 | 0.010 | 1.000 |  |  |  |  |  |  | 147.934 |
| Inverse variance weighted | 0.022 | 0.011 | 0.043 |  |  | 750.172 | 213 | 5.88x10<sup>-61</sup> | 71.607 | 147.934 |
| Simple mode | -0.025 | 0.018 | 0.168 |  |  |  |  |  |  | 147.934 |
| Weighted mode | -0.001 | 0.011 | 0.938 |  |  |  |  |  |  | 147.934 |
| **Univariable MR Analysis — Exposure: RETIC (210 SNPs), Outcome: FI** |  |  |  |  |  |  |  |  |  |  |
| MR Egger | -0.001 | 0.017 | 0.931 | 0.001 | 0.024 | 761.249 | 208 | 4.25x10<sup>-64</sup> | 72.676 | 316.202 |
| Weighted median | 0.011 | 0.010 | 0.259 |  |  |  |  |  |  | 316.202 |
| Inverse variance weighted | 0.031 | 0.010 | 0.002 |  |  | 780.233 | 209 | 7.81x10<sup>-67</sup> | 73.213 | 316.202 |
| Simple mode | 0.008 | 0.021 | 0.694 |  |  |  |  |  |  | 316.202 |
| Weighted mode | 0.003 | 0.012 | 0.768 |  |  |  |  |  |  | 316.202 |

## Supplementary File 5 — Bidirectional MR analyses assessing the effect of Hb on HbA1c

### Description of Different Figure Types

A. Scatter plot showing the single nucleotide polymorphisms (SNPs) associated with Hb against SNPs associated with HbA1c as the outcome (vertical and horizontal black lines around points show 95% confidence intervals (CI)) for five different Mendelian Randomization (MR) association tests.
B. Funnel plot of the effect size against the inverse of the standard error for each SNP.

Exposure: Hb, Outcome: HbA1c

Figures of Scatter Plot of MR Methods (A) and Funnel Plot (B)

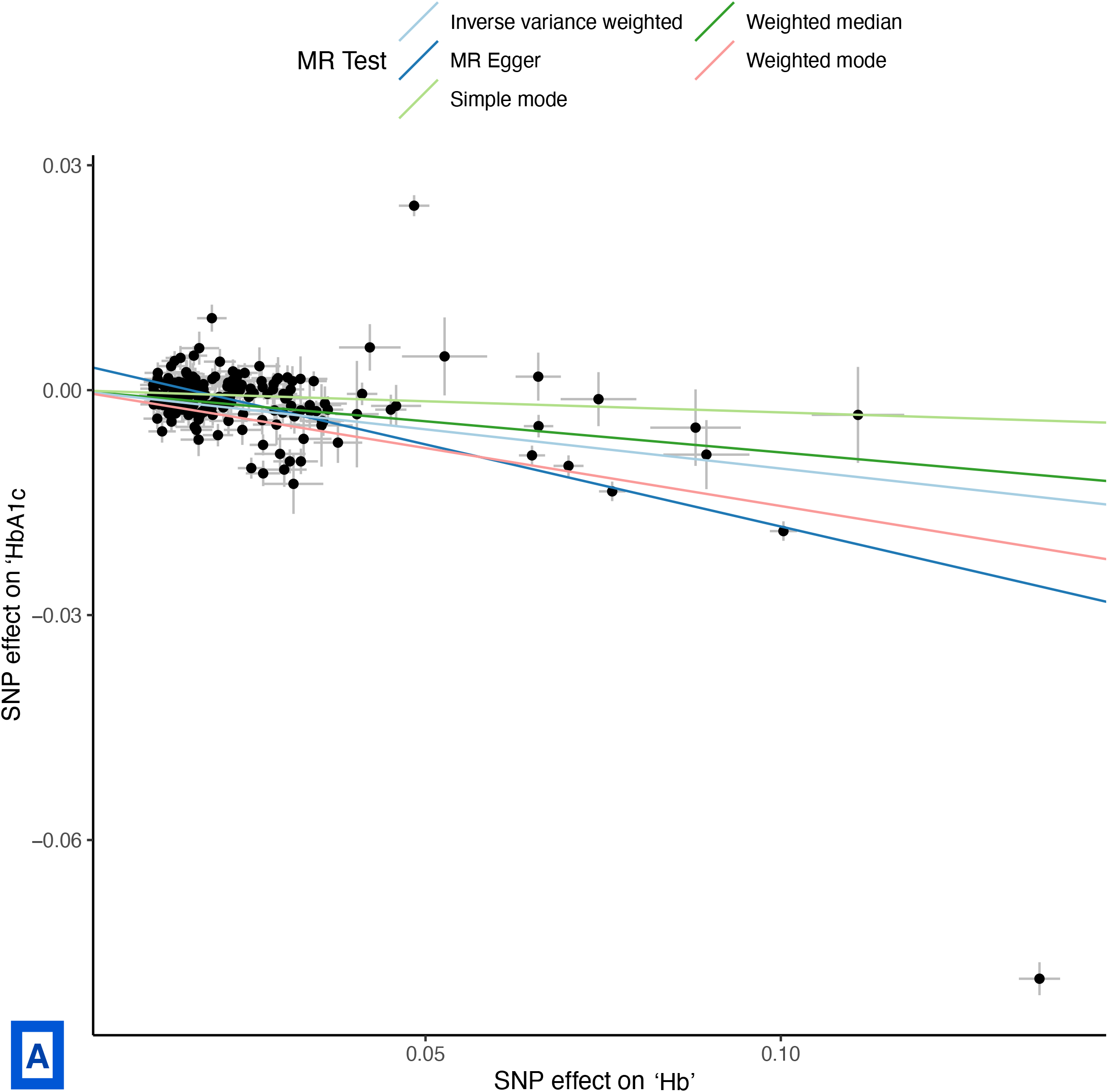
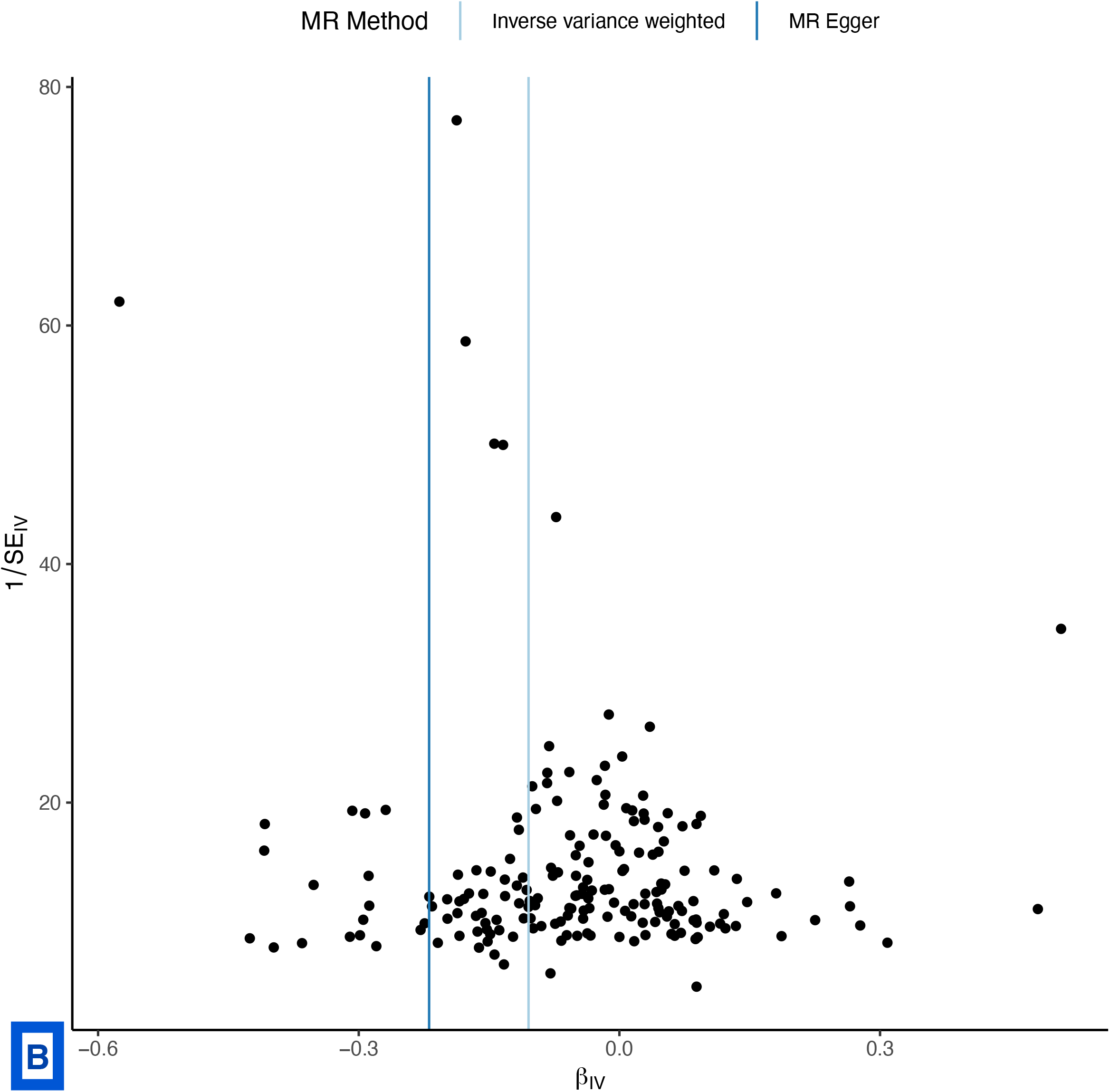

Exposure: HbA1c, Outcome: Hb

Figures of Scatter Plot of MR Methods (A) and Funnel Plot (B)

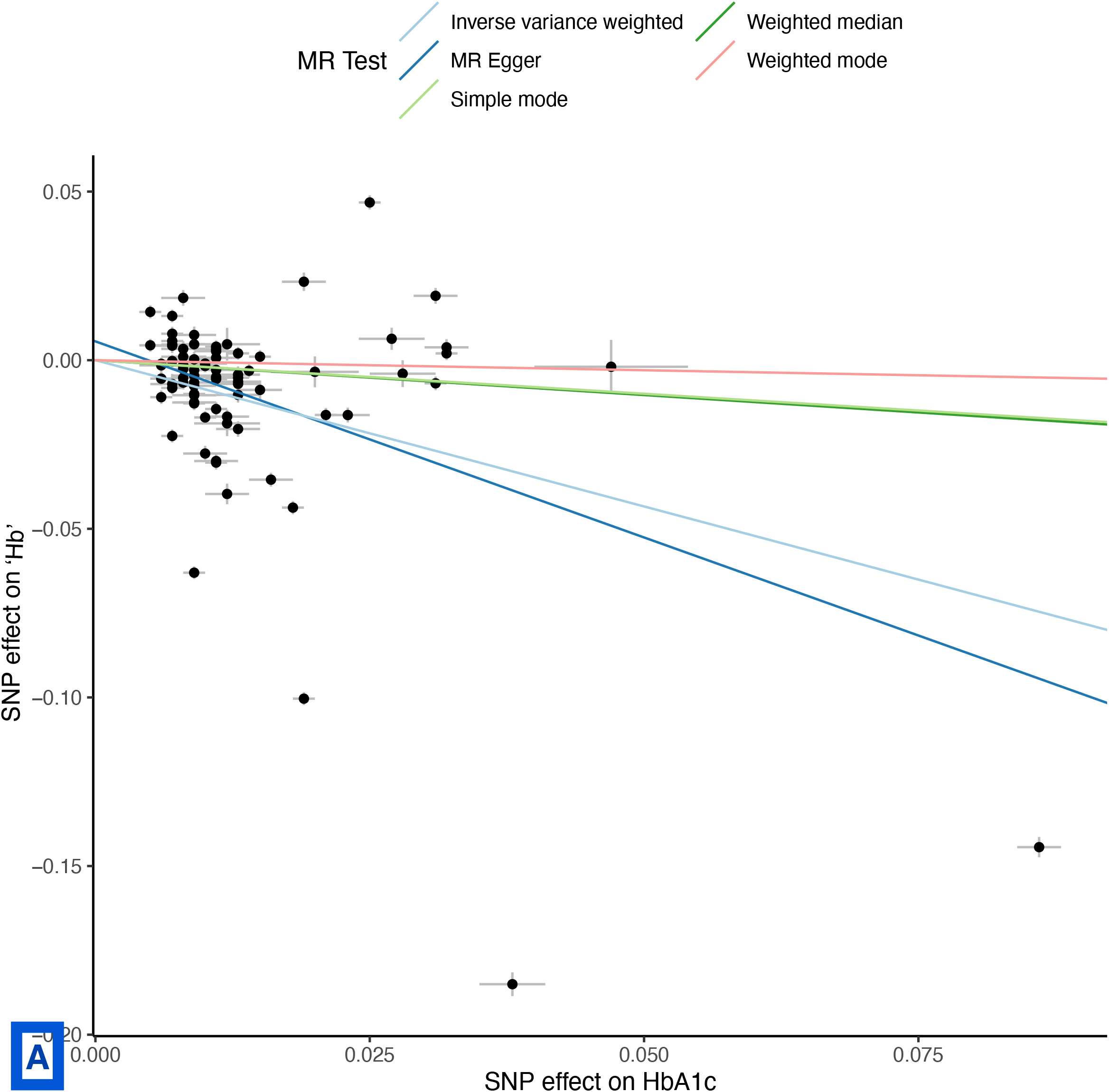
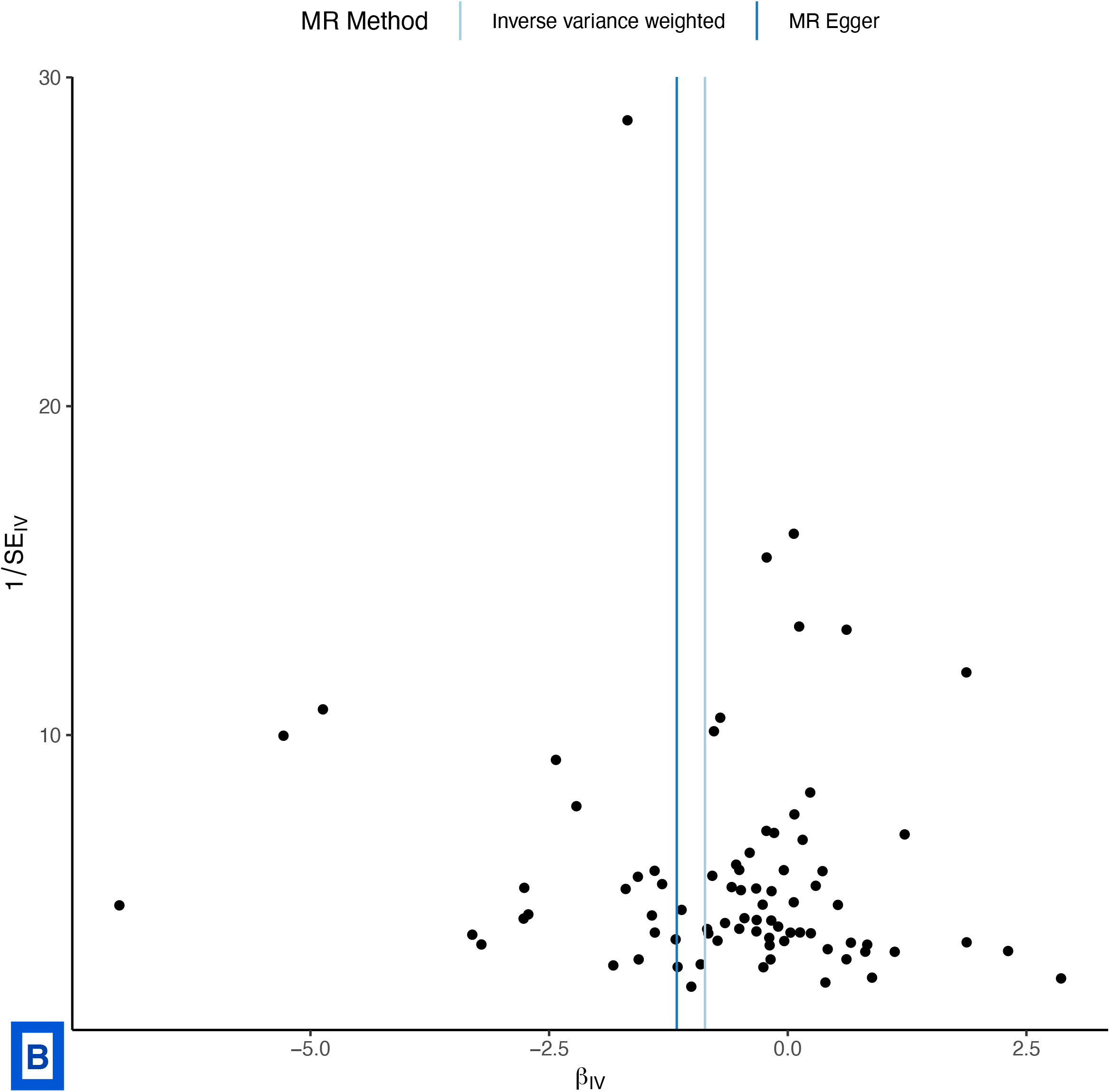

## Supplemental File 6: Baseline characteristics for participants of observational study and descriptive statistics

**Table:** Baseline characteristics for participants of observational study.

| Characteristic | Healthy Participants,<br>N = 6,504<sup>1</sup> | Participants with<br>Pre-T2D, N = 1,096<sup>1</sup> | p-value<sup>2</sup> |
| --- | --- | --- | --- |
| Sex |  |  | 0.076 |
| Female | 3,147 (48%) | 562 (51%) |  |
| Male | 3,357 (52%) | 534 (49%) |  |
| Age (years) | 50 (41, 58) | 56 (49, 61) | <0.001 |
| HbA1c (%) | 5.40 (5.20, 5.60) | 6.10 (6.00, 6.20) | <0.001 |
| Fasting Glucose (mmol/L) | 5.10 (4.70, 5.50) | 5.70 (5.20, 6.20) | <0.001 |
| Triglyceride (mmol/L) | 1.16 (0.85, 1.68) | 1.33 (0.99, 1.92) | <0.001 |
| HDL Cholesterol (mmol/L) | 1.29 (1.07, 1.57) | 1.19 (1.01, 1.43) | <0.001 |
| Hemoglobin (g/L) | 145 (136, 154) | 144 (134, 153) | <0.001 |
| Triglyceride Glucose Index | 8.46 (8.12, 8.85) | 8.71 (8.36, 9.08) | <0.001 |
| Predicted A1c, adjusted for<br>age, sex (%) | 5.04 (4.97, 5.11) | 5.14 (5.06, 5.23) | <0.001 |
| Glycation Gap<sup>3</sup> | 0.36 (0.12, 0.57) | 1.01 (0.89, 1.13) | <0.001 |
| <sup>1</sup> Median (IQR); n (%) |  |  |  |
| <sup>2</sup> Wilcoxon rank sum test; Pearson's Chi-squared test |  |  |  |
| <sup>3</sup> Actual HbA1c – Predicted HbA1c |  |  |  |

**Table:** Linear Regression for Triglyceride Glucose Index (exposure) on Glycation Gap (outcome)*

| Population (n) | Beta | Standard error | y-intercept | p-value |
| --- | --- | --- | --- | --- |
| Pre-T2D (1096) | -0.087 | 0.009 | 1.77 | <0.0001 |
| Healthy (6504) | 0.023 | 0.007 | 0.14 | <0.0001 |
| HbA1c < 5%<br>(681) | 0.071 | 0.01 | 0.41 | <0.0001 |
| HbA1c 5 – 5.4%<br>(2948) | 0.047 | 0.005 | 0.6 | <0.0001 |
| HbA1c 5.5 –<br>5.9%<br>(2875) | 0.054 | 0.006 | 1.06 | <0.0001 |

\*calculated as Actual HbA1c – Predicted HbA1c using model adjusted for age, sex (18)

## Supplementary File 7

Link: https://www.dropbox.com/s/827zwgfwszefe36/SF8_FP_LOO_Export_All_Analyses.xlsx?dl=0

## Notes

### Competing Interest Statement

The authors have declared no competing interest.

### Author Declarations

For our primary analysis we undertook bidirectional inverse variance weighted (IVW) MR with FI as exposure (Supplementary File 3) and Hb, red cell count (RCC), reticulocyte count (RETIC) as outcomes. A p value of <0.05 was considered significant for primary and secondary analyses. We followed the recently published STROBE-MR reporting guidelines (Checklist in Supplementary File 2)(26). As we used publicly available summary statistics from GWAS, we did not seek institutional approval. Informed consent was obtained from the investigators from each participant in the original study Observational Study We received institutional approval from UHN research ethics board for the observational study. As we analyzed anonymized data, we did not obtain consent from individual patients.

## References

1. Selvin E. Hemoglobin A 1c-Using Epidemiology to Guide Medical Practice: Kelly West Award Lecture 2020. Diabetes Care [Internet]. Diabetes Care; 2021 [cited 2022 Aug 16];44:2197–204. Available from: https://pubmed.ncbi.nlm.nih.gov/34548283/

2. WHO Global Report. Global Report on Diabetes. Isbn [Internet]. 2016 [cited 2022 Aug 21];978:11. Available from: http://www.who.int/about/licensing/copyright_form/index.html %0A http://www.who.int/about/licensing/copyright_form/index.html %0A http://www.who.int/about/licensing/copyright_form/index.html %0A https://apps.who.int/iris/handle/10665/204871 %0A http://www.who.int

3. Tabák AG, Jokela M, Akbaraly TN, Brunner EJ, Kivimäki M, Witte DR. Trajectories of glycaemia, insulin sensitivity, and insulin secretion before diagnosis of type 2 diabetes: an analysis from the Whitehall II study. The Lancet. Elsevier B.V.; 2009;373:2215–21.

4. Aminian A, Vidal J, Salminen P, Still CD, Hanipah ZN, Sharma G, et al. Late Relapse of Diabetes After Bariatric Surgery: Not Rare, but Not a Failure. Diabetes Care [Internet]. Diabetes Care; 2020 [cited 2022 Aug 16];43:534–40. Available from: https://pubmed.ncbi.nlm.nih.gov/31974105/

5. Lean MEJ, Leslie WS, Barnes AC, Brosnahan N, Thom G, McCombie L, et al. Durability of a primary care-led weight-management intervention for remission of type 2 diabetes: 2-year results of the DiRECT open-label, cluster-randomised trial. Lancet Diabetes Endocrinol [Internet]. Lancet Diabetes Endocrinol; 2019 [cited 2022 Aug 16];7:344–55. Available from: https://pubmed.ncbi.nlm.nih.gov/30852132/

6. Nathan DM, Barrett-Connor E, Crandall JP, Edelstein SL, Goldberg RB, Horton ES, et al. Long-term Effects of Lifestyle Intervention or Metformin on Diabetes Development and Microvascular Complications: the DPP Outcomes Study. Lancet Diabetes Endocrinol [Internet]. NIH Public Access; 2015 [cited 2022 Aug 16];3:866. Available from: /pmc/articles/PMC4623946/

7. Berard LD, Siemens R, Pharm B, Woo V. 2018 Clinicsal Practice Guidelines. 2018 [cited 2022 Aug 16]; Available from: https://doi.org/10.1016/j.jcjd.2017.10.007

8. Davies MJ, D’Alessio DA, Fradkin J, Kernan WN, Mathieu C, Mingrone G, et al. Management of hyperglycaemia in type 2 diabetes, 2018. A consensus report by the American Diabetes Association (ADA) and the European Association for the Study of Diabetes (EASD). Diabetologia [Internet]. Diabetologia; 2018 [cited 2022 Aug 16];61:2461–98. Available from: https://pubmed.ncbi.nlm.nih.gov/30288571/

9. Riddle MC, Cefalu WT, Evans PH, Gerstein HC, Nauck MA, Oh WK, et al. Consensus Report: Definition and Interpretation of Remission in Type 2 Diabetes. Diabetes Care [Internet]. Diabetes Care; 2021 [cited 2022 Aug 16];44:2438–44. Available from: https://pubmed.ncbi.nlm.nih.gov/34462270/

10. Reed J, Bain S, Kanamarlapudi V. A review of current trends with type 2 diabetes epidemiology, aetiology, pathogenesis, treatments and future perspectives. Diabetes Metab Syndr Obes. Dove Medical Press Ltd; 2021. page 3567–602.

11. Lacy ME, Wellenius GA, Sumner AE, Correa A, Carnethon MR, Liem RI, et al. Association of Sickle Cell Trait With Hemoglobin A1c in African Americans. JAMA [Internet]. American Medical Association; 2017 [cited 2022 Aug 16];317:507–15. Available from: https://jamanetwork.com/journals/jama/fullarticle/2600468

12. Fizelova M, Stančáková A, Lorenzo C, Haffner SM, Cederberg H, Kuusisto J, et al. Glycated Hemoglobin Levels Are Mostly Dependent on Nonglycemic Parameters in 9398 Finnish Men Without Diabetes. J Clin Endocrinol Metab [Internet]. Oxford Academic; 2015 [cited 2022 Aug 16];100:1989–96. Available from: https://academic.oup.com/jcem/article/100/5/1989/2829717

13. Jansen H, Stolk RP, Nolte IM, Kema IP, Wolffenbuttel BHR, Snieder H. Determinants of HbA1c in nondiabetic Dutch adults: Genetic loci and clinical and lifestyle parameters, and their interactions in the lifelines cohort study. J Intern Med. 2013;273:283–93.

14. Barazzoni R, Cappellari GG, Semolic A, Chendi E, Ius M, Situlin R, et al. The Association between Hematological Parameters and Insulin Resistance Is Modified by Body Mass Index – Results from the North-East Italy MoMa Population Study. PLoS One [Internet]. Public Library of Science; 2014 [cited 2022 Aug 16];9:e101590. Available from: https://journals.plos.org/plosone/article?id=10.1371/journal.pone.0101590

15. Woo M, Hawkins M. Beyond Erythropoiesis: Emerging Metabolic Roles of Erythropoietin. Diabetes [Internet]. American Diabetes Association; 2014 [cited 2022 Aug 16];63:2229–31. Available from: https://diabetesjournals.org/diabetes/article/63/7/2229/34347/Beyond-Erythropoiesis-Emerging-Metabolic-Roles-of

16. Barbieri M, Ragno E, Benvenuti E, Zito GA, Corsi A, Ferrucci L, et al. New aspects of the insulin resistance syndrome: impact on haematological parameters.

17. Hemani G, Zheng J, Elsworth B, Wade KH, Haberland V, Baird D, et al. The MR-Base platform supports systematic causal inference across the human phenome. 2018; Available from: https://doi.org/10.7554/eLife.34408.001

18. Simental-Mendía LE, Rodríguez-Morán M, Guerrero-Romero F. The product of fasting glucose and triglycerides as surrogate for identifying insulin resistance in apparently healthy subjects. Metab Syndr Relat Disord [Internet]. Metab Syndr Relat Disord; 2008 [cited 2022 Oct 23];6:299–304. Available from: https://pubmed.ncbi.nlm.nih.gov/19067533/

19. Chen J, Spracklen CN, Marenne G, Varshney A, Corbin LJ, Luan J, et al. The trans-ancestral genomic architecture of glycemic traits. Nat Genet [Internet]. Nat Genet; 2021 [cited 2022 Aug 27];53:840–60. Available from: https://pubmed.ncbi.nlm.nih.gov/34059833/

20. Vuckovic D, Bao EL, Akbari P, Lareau CA, Mousas A, Jiang T, et al. The Polygenic and Monogenic Basis of Blood Traits and Diseases. Cell [Internet]. Cell; 2020 [cited 2022 Aug 27];182:1214-1231.e11. Available from: https://pubmed.ncbi.nlm.nih.gov/32888494/

21. Chen MH, Raffield LM, Mousas A, Sakaue S, Huffman JE, Moscati A, et al. Trans-ethnic and Ancestry-Specific Blood-Cell Genetics in 746,667 Individuals from 5 Global Populations. Cell [Internet]. Cell; 2020 [cited 2022 Aug 27];182:1198-1213.e14. Available from: https://pubmed.ncbi.nlm.nih.gov/32888493/

22. Mbatchou J, Barnard L, Backman J, Marcketta A, Kosmicki JA, Ziyatdinov A, et al. Computationally efficient whole-genome regression for quantitative and binary traits. Nat Genet [Internet]. Nat Genet; 2021 [cited 2022 Aug 27];53:1097–103. Available from: https://pubmed.ncbi.nlm.nih.gov/34017140/

23. Xue A, Wu Y, Zhu Z, Zhang F, Kemper KE, Zheng Z, et al. Genome-wide association analyses identify 143 risk variants and putative regulatory mechanisms for type 2 diabetes. Nat Commun [Internet]. Nat Commun; 2018 [cited 2022 Jul 14];9. Available from: https://pubmed.ncbi.nlm.nih.gov/30054458/

24. Scott RA, Lagou V, Welch RP, Wheeler E, Montasser ME, Luan J, et al. Large-scale association analyses identify new loci influencing glycemic traits and provide insight into the underlying biological pathways. Nat Genet [Internet]. Nat Genet; 2012 [cited 2022 Jul 14];44:991–1005. Available from: https://pubmed.ncbi.nlm.nih.gov/22885924/

25. Pulit SL, Stoneman C, Morris AP, Wood AR, Glastonbury CA, Tyrrell J, et al. Meta-analysis of genome-wide association studies for body fat distribution in 694 649 individuals of European ancestry. Hum Mol Genet [Internet]. Hum Mol Genet; 2019 [cited 2022 Jul 14];28:166–74. Available from: https://pubmed.ncbi.nlm.nih.gov/30239722/

26. Skrivankova VW, Richmond RC, Woolf BAR, Davies NM, Swanson SA, Vanderweele TJ, et al. Strengthening the reporting of observational studies in epidemiology using mendelian randomisation (STROBE-MR): explanation and elaboration. BMJ [Internet]. British Medical Journal Publishing Group; 2021 [cited 2022 Jul 16];375. Available from: https://www.bmj.com/content/375/bmj.n2233

27. Bowden J, Smith GD, Burgess S. Mendelian randomization with invalid instruments: effect estimation and bias detection through Egger regression. Int J Epidemiol [Internet]. Int J Epidemiol; 2015 [cited 2022 Jul 15];44:512–25. Available from: https://pubmed.ncbi.nlm.nih.gov/26050253/

28. Bowden J, Davey Smith G, Haycock PC, Burgess S. Consistent Estimation in Mendelian Randomization with Some Invalid Instruments Using a Weighted Median Estimator. Genet Epidemiol [Internet]. Genet Epidemiol; 2016 [cited 2022 Jul 15];40:304–14. Available from: https://pubmed.ncbi.nlm.nih.gov/27061298/

29. Hartwig FP, Smith GD, Bowden J. Robust inference in summary data Mendelian randomization via the zero modal pleiotropy assumption. Int J Epidemiol [Internet]. Int J Epidemiol; 2017 [cited 2022 Jul 15];46:1985–98. Available from: https://pubmed.ncbi.nlm.nih.gov/29040600/

30. Sanderson E, Spiller W, Bowden J. Testing and correcting for weak and pleiotropic instruments in two-sample multivariable Mendelian randomization. Stat Med [Internet]. Stat Med; 2021 [cited 2022 Jul 1];40:5434–52. Available from: https://pubmed.ncbi.nlm.nih.gov/34338327/

31. Sadreev II, Elsworth BL, Mitchell RE, Paternoster L, Sanderson E, Davies NM, et al. Navigating sample overlap, winner’s curse and weak instrument bias in Mendelian randomization studies using the UK Biobank. medRxiv [Internet]. Cold Spring Harbor Laboratory Press; 2021 [cited 2022 Jul 14];2021.06.28.21259622. Available from: https://www.medrxiv.org/content/10.1101/2021.06.28.21259622v1

32. Sanderson E, Davey Smith G, Windmeijer F, Bowden J. An examination of multivariable Mendelian randomization in the single-sample and two-sample summary data settings. Int J Epidemiol [Internet]. Oxford Academic; 2019 [cited 2022 Jul 1];48:713–27. Available from: https://academic.oup.com/ije/article/48/3/713/5238110

33. Inzucchi SE, Zinman B, Fitchett D, Wanner C, Ferrannini E, Schumacher M, et al. How Does Empagliflozin Reduce Cardiovascular Mortality? Insights From a Mediation Analysis of the EMPA-REG OUTCOME Trial. Diabetes Care [Internet]. Diabetes Care; 2018 [cited 2022 Oct 23];41:356–63. Available from: https://pubmed.ncbi.nlm.nih.gov/29203583/

34. Ratajczak J, Zhang Q, Pertusini E, Wojczyk BS, Wasik MA, Ratajczak MZ. The role of insulin (INS) and insulin-like growth factor-I (IGF-I) in regulating human erythropoiesis. Studies in vitro under serum-free conditions--comparison to other cytokines and growth factors. Leukemia [Internet]. Leukemia; 1998 [cited 2022 Oct 23];12:371–81. Available from: https://pubmed.ncbi.nlm.nih.gov/9529132/

35. Efron B, Tibshirani R. Improvements on Cross-Validation: The .632+ Bootstrap Method. J Am Stat Assoc. JSTOR; 1997;92:548.

36. Semple RK, Sleigh A, Murgatroyd PR, Adams CA, Bluck L, Jackson S, et al. Postreceptor insulin resistance contributes to human dyslipidemia and hepatic steatosis. J Clin Invest [Internet]. J Clin Invest; 2009 [cited 2022 Aug 30];119:315–22. Available from: https://pubmed.ncbi.nlm.nih.gov/19164855/

37. Ratajczak J, Zhang Q, Pertusini E, Wojczyk BS, Wasik MA, Ratajczak MZ. The role of insulin (INS) and insulin-like growth factor-I (IGF-I) in regulating human erythropoiesis. Studies in vitro under serum-free conditions--comparison to other cytokines and growth factors. Leukemia [Internet]. Leukemia; 1998 [cited 2022 Aug 30];12:371–81. Available from: https://pubmed.ncbi.nlm.nih.gov/9529132/

38. Griffin SJ, Borch-Johnsen K, Davies MJ, Khunti K, Rutten GE, Sandbæk A, et al. Effect of early intensive multifactorial therapy on 5-year cardiovascular outcomes in individuals with type 2 diabetes detected by screening (ADDITION-Europe): a cluster-randomised trial. Lancet [Internet]. Lancet; 2011 [cited 2022 Aug 27];378:156–67. Available from: https://pubmed.ncbi.nlm.nih.gov/21705063/

39. Sarnowski C, Leong A, Raffield LM, Wu P, de Vries PS, DiCorpo D, et al. Impact of Rare and Common Genetic Variants on Diabetes Diagnosis by Hemoglobin A1c in Multi-Ancestry Cohorts: The Trans-Omics for Precision Medicine Program. Am J Hum Genet [Internet]. Am J Hum Genet; 2019 [cited 2022 Aug 30];105:706–18. Available from: https://pubmed.ncbi.nlm.nih.gov/31564435/

40. Chai JF, Kao SL, Wang C, Jun-Yu Lim V, Khor IW, Dou J, et al. Genome-Wide Association for HbA1c in Malay Identified Deletion on SLC4A1 that Influences HbA1c Independent of Glycemia. J Clin Endocrinol Metab [Internet]. J Clin Endocrinol Metab; 2020 [cited 2022 Aug 30];105:3854–64. Available from: https://pubmed.ncbi.nlm.nih.gov/32936915/

41. Wu W-C, Lacy ME, Correa A, Carnethon M, Reiner AP, Eaton CB, et al. Association Between Hemoglobin A1c and Glycemia in African Americans with and without Sickle Cell trait and Whites, Results from CARDIA and the Jackson Heart Study. J Diabetes Treat [Internet]. NIH Public Access; 2018 [cited 2022 Aug 30];3. Available from: /pmc/articles/PMC8939875/

42. Paterson AD. HbA1c for type 2 diabetes diagnosis in Africans and African Americans: Personalized medicine NOW! PLoS Med [Internet]. Public Library of Science; 2017 [cited 2022 Aug 30];14:e1002384. Available from: https://journals.plos.org/plosmedicine/article?id=10.1371/journal.pmed.1002384

43. Lacy ME, Wellenius GA, Sumner AE, Correa A, Carnethon MR, Liem RI, et al. Association of Sickle Cell Trait With Hemoglobin A1c in African Americans. JAMA [Internet]. American Medical Association; 2017 [cited 2022 Aug 30];317:507–15. Available from: https://jamanetwork.com/journals/jama/fullarticle/2600468

44. Lagou V, Mägi R, Hottenga JJ, Grallert H, Perry JRB, Bouatia-Naji N, et al. Sex-dimorphic genetic effects and novel loci for fasting glucose and insulin variability. Nature Communications 2021 12:1 [Internet]. Nature Publishing Group; 2021 [cited 2022 Aug 29];12:1–18. Available from: https://www.nature.com/articles/s41467-020-19366-9

## References

1. Skrivankova VW, Richmond RC, Woolf BAR, Yarmolinsky J, Davies NM, Swanson SA, et al. Strengthening the Reporting of Observational Studies in Epidemiology using Mendelian Randomization (STROBE-MR) Statement. JAMA. 2021;under review.

2. Skrivankova VW, Richmond RC, Woolf BAR, Davies NM, Swanson SA, VanderWeele TJ, et al. Strengthening the Reporting of Observational Studies in Epidemiology using Mendelian Randomisation (STROBE-MR): Explanation and Elaboration. BMJ. 2021;375:n2233.

